# Weight Change Between Pregnancies and Mortality Over 50 Years of Follow-up

**DOI:** 10.1101/2025.09.22.25336340

**Authors:** Yajnaseni Chakraborti, Sunni L. Mumford, Edwina H. Yeung, Katherine L. Grantz, Pauline Mendola, James L. Mills, Ellen C. Caniglia, Colleen M. Brensinger, Cuilin Zhang, Enrique F. Schisterman, Stefanie N. Hinkle

## Abstract

**Objective:** To evaluate the link between postpartum weight retention (PPWR) and long-term mortality.

**Methods:** In this secondary analysis of 8165 women with ≥2 pregnancies in the Collaborative Perinatal Project(CPP), U.S., 1959–1966, with vital status follow-up through 2016, we used interconception weight change (ICWC) and interpregnancy weight change (IPWC) as proxies of PPWR. ICWC was defined as the difference between self-reported pre-pregnancy weights from the 1^st^ and 2^nd^ CPP pregnancies, while IPWC was the difference between weight recorded at delivery of the 1^st^ and pre-pregnancy weight of the 2^nd^ CPP pregnancies. All-cause and cause-specific mortality models were adjusted for sociodemographic, behavioral, clinical, and pregnancy-related characteristics from the 1^st^ CPP pregnancy.

**Results:** Compared to women with ICWC >0 to 1.8 kg (quintile 3), those with ICWC > –1.4 to 0 kg (quintile 2) had a lower risk of all-cause mortality [aHR (95% CI): 0.85 (0.76–0.96)], with the most notable reduction observed in diabetes-related risk of mortality [aHR (95% CI): 0.45 (0.23-0.87)]. Lower quintiles of IPWC (i.e., greater weight loss) were suggestive of reduced all-cause and cause-specific risk of mortality, though the estimates were not statistically significant.

**Conclusions:** Minimizing PPWR was linked to reduced mortality risk over 50 years of follow-up.

## Introduction

Over the past three decades, the prevalence of overweight and obesity among women of reproductive age has steadily increased(1–3). Currently, more than one in four women between the ages of 20 and 39 in the U.S. are living with overweight (27.5%) or obesity (36.8%)(1, 2, 4). For many women, weight gained during pregnancy may contribute to the development of overweight or obesity(5). In fact, the 5-incidence of obesity is twice as high among women who have had a child compared to those who have never given birth(6). A comparable pattern of risk was reported in a separate study investigating childbearing and weight gain over 10 years(7). A probable mechanism underlying this risk of obesity associated with parity is postpartum weight retention (PPWR), which refers to the weight a person retains after giving birth, in addition to the weight gained during pregnancy.

Only about one in four women returns to their pre-pregnancy weight by 1-year postpartum(8). For instance, a U.S.-based prospective cohort study by Endres et al. (2015), found that approximately 75% of participants weighed more one year after giving birth than they did before pregnancy—at 1-year postpartum, 47.4% retained more than 10 pounds and 24.2% retained more than 20 pounds. Further, among those with a normal pre-pregnancy body mass index (BMI), 33% were living with overweight or obesity at 1-year postpartum(8). Legro et al. (2020) reported similar findings in their study of first-time mothers in Pennsylvania, where 23.7% retained between 1 and 9 pounds, and 23.9% retained 10 or more pounds at 1-year postpartum. Among those with a normal pre-pregnancy BMI, 12% were living with overweight or obesity at 1-year postpartum(9).

Indeed, gestational weight gain (GWG) is a likely contributor to PPWR(5, 10–12), with studies showing associations between excessive GWG and greater overall and abdominal adiposity even 8 to 15 years postpartum(13). However, GWG is not the only factor influencing PPWR; other contributors, such as lifestyle behaviors and breastfeeding, may also affect whether weight is retained postpartum(12, 14–21). Given that multiple factors contribute to PPWR, it is important to examine its impact on health as a distinct exposure, rather than viewing it solely as a consequence of GWG(10). In fact, one study found that post-partum weight trajectories were associated with excess weight and increased adiposity risk at 3 years postpartum, even after adjusting for GWG(12).

Additionally, the implications of PPWR on cardiometabolic health are well documented with adverse cardiometabolic profiles often emerging within the first year postpartum(22), and elevated risks of pre-diabetes or diabetes(23), poor cardiometabolic profiles(12, 23), as well as hypertensive and cardiovascular disorders(24) continuing to manifest at 3-(12, 23), 5-(23), and 16-years(24) postpartum.

Given the evidence of sustained alterations in weight and cardiometabolic parameters, it is reasonable to hypothesize that PPWR may be related to long-term mortality, potentially independent of pre-pregnancy BMI and gestational weight gain, yet this remains unexplored. To evaluate this hypothesis, we estimated the risk of all-cause mortality over 50 years of follow-up post-pregnancy, in relation to two analytic exposures: 1) interconception weight change, and 2) interpregnancy weight change, which are operational proxies for PPWR capturing maternal weight dynamics across distinct reproductive intervals. Additionally, given the existing evidence linking PPWR to cardiovascular and metabolic disorders(25, 26), we also examined the cardiovascular, diabetes, and kidney-specific mortality risks associated with these two exposures.

## Methods

This study is a secondary analysis of data collected from the Collaborative Perinatal Project (CPP) and the CPP Mortality Linkage Study. The CPP was a multi-center observational prospective cohort study conducted in the U.S. between 1959 and 1966 to assess the effects of pregnancy and perinatal complications on birth and child outcomes(27–32). A total of 48,917 pregnant women were enrolled at their first prenatal visit, and were followed across multiple pregnancies, resulting in 58,760 registered pregnancies. The mortality status of this cohort as of Dec 31, 2016, was later determined through the CPP Mortality Linkage study(33), to explore the long-term implications of gestational exposures. This analysis utilizes data from the first two pregnancies for participants with two or more CPP pregnancies (n = 8165). It is important to note that the 1^st^ pregnancy registered within the CPP was not necessarily an individual’s 1st pregnancy overall. For the purposes of this study, we have used the term “1^st^ pregnancy” to refer specifically to the 1^st^ pregnancy recorded in the CPP.

There were no specific regulations for approving research involving human subjects at the time the CPP began in the late 1950s, yet general informed consent for participation was obtained. IRB approval for the CPP Mortality Linkage Study was obtained by the *Eunice Kennedy Shriver* National Institute of Child Health and Human Development (IRB00011862, approved in 2015) and the Emmes Corporation (IRB00000879, approved in 2013)—the two organizations responsible for abstracting identifying information from historic CPP records and facilitating the data linkages to the National Death Index (NDI) and the Social Security Death Master File (SSDMF). Mortality data were linked through December 31, 2016. The University of Pennsylvania IRB entered into a reliance agreement with the NICHD IRB for oversight of this study.

### Exposures

Participants self-reported their pre-pregnancy weight at their first prenatal visit for each CPP pregnancy. Participants’ weight at each prenatal visit and delivery admission was abstracted from medical records. This analysis conceptualized PPWR using two different weight exposures based on the participants’ consecutive 1^st^ and 2^nd^ CPP pregnancies (Figure 1). The primary exposure was interconception weight change (ICWC), which represented the overall weight change between the 1^st^ and 2^nd^ CPP pregnancies. Specifically, ICWC was calculated as the difference between pre-pregnancy weight of the 2^nd^ and 1^st^ pregnancies. We also examined a secondary exposure, interpregnancy weight change (IPWC), which focused solely on weight change in the interpregnancy interval (IPI). Specifically, IPWC was calculated as the difference between pre-pregnancy weight of the 2^nd^ CPP pregnancy and weight recorded at admission for delivery of the 1^st^ CPP pregnancy. We preprocessed height and weight measurements by identifying and setting implausible values to missing based on predefined thresholds (detailed specifications in Appendix A).

**Figure 1:**
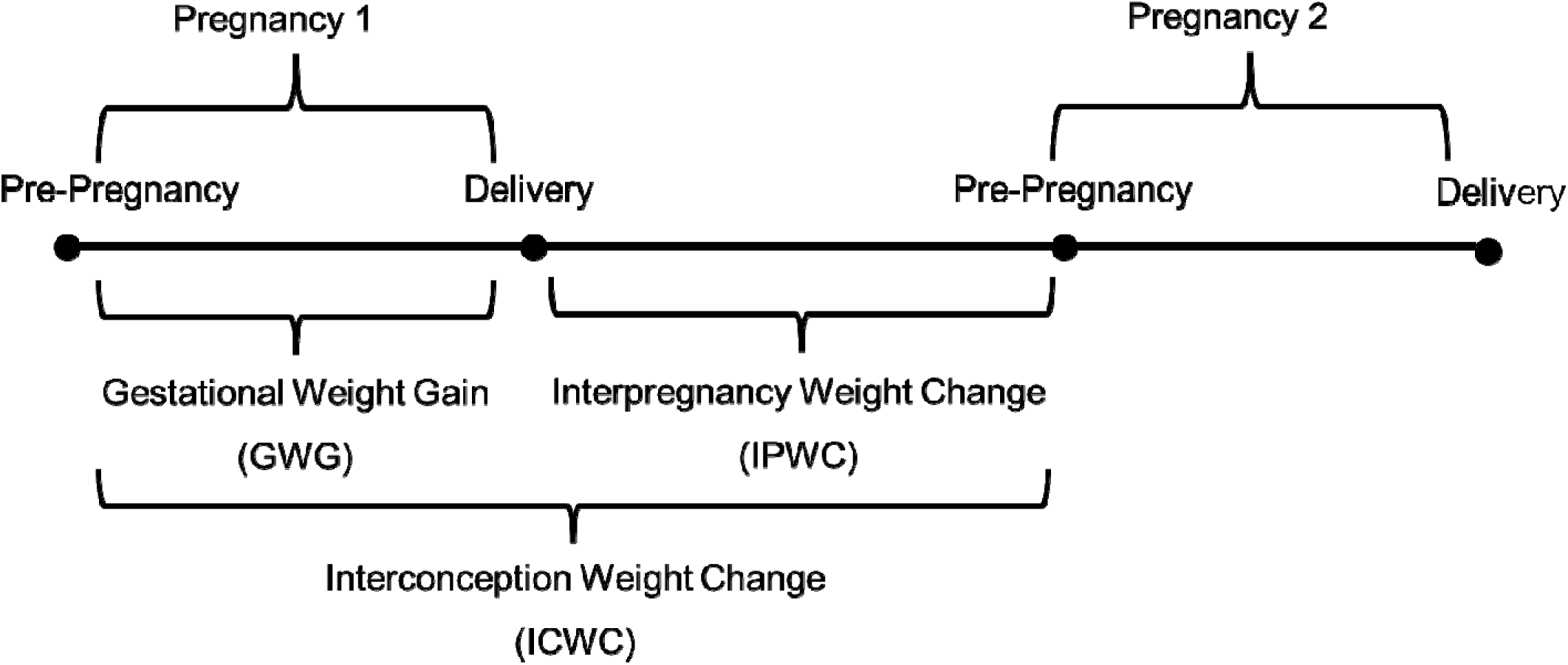
Exposures and timeline of measurement

### Outcomes

The primary outcome of interest was time to all-cause mortality, defined as the time from the start of follow up, to the date of death or Dec 31, 2016, whichever occurred first. Follow up started at the date of the last menstrual period (LMP) of the participant’s 2^nd^ CPP pregnancy. All-cause mortality was ascertained through the National Death Index (NDI); deaths not identified in the NDI were captured using the Social Security Death Master File (SSDMF). The NDI includes all U.S. death records from 1979 onward, with matches identified through probabilistic linkage. Although the SSDMF includes deaths dating back to 1958, the NDI is considered a more accurate and comprehensive source of mortality and also includes cause of death(33, 34). Overall, 89% of the cohort’s death information was sourced from the NDI. Given the link between PPWR and cardiometabolic health(12, 23), secondary outcomes considered in this study were time to cause-specific mortality related to cardiovascular disease (I00-I78, I80-I99), diabetes (E10-E14), and kidney conditions (N00-N07, N17-N19, N25-N27) where the underlying cause of death was coded using ICD-10 (International Classification of Diseases, 10^th^ Revision) and comparable ICD-9 codes. Some misclassification of cause of death is possible. Because only approximately 3% of deaths occurred before 1979(33), the impact of missing cause-of-death information is expected to be minimal.

### Covariates

Covariates were reported by participants during the first prenatal visit of their 1^st^ CPP pregnancy, and included information on age, frequency of smoking, race, marital status, income, education, number of prior pregnancies, hypertensive disorders of pregnancy(35), diabetes prior to pregnancy, prior cardiovascular diseases, prior respiratory diseases, prior renal disease, prior neurological conditions, prior cancer/tumor status, study site, pre-pregnancy weight, and height. Race was conceptualized as a social determinant of health in this study. Body mass index (BMI) (kg/m²) was calculated using self-reported height and pre-pregnancy weight. Additional 1^st^ CPP pregnancy variables of interest included in this study were pregnancy plurality, gestational age at delivery, pregnancy outcomes, GWG, and the interconception and interpregnancy intervals. Details on how these variables were derived and incorporated at different analytical steps of the study are provided in Appendix B—Supplementary Table 1.

### Statistical Analysis

Post data cleaning and processing, the participant characteristics were summarized for each quintile of ICWC. Missing or implausible exposure, covariate and outcome data were imputed via multiple imputation using chained equations. Under the Missing at Random assumption, 10 imputed datasets were generated for estimating pooled hazard ratios and the respective 95% Confidence Interval (CI). Multiple imputation was conducted using the R package *mice*(36). All other data cleaning, processing and analyses were conducted using SAS.

The study sample was a subset of the CPP cohort with at least two pregnancies in the CPP. Thus, to overcome the potential bias due to this inherent selection process and to ensure that the pooled hazard ratio estimates are generalizable to the entire CPP target-population, inverse probability weights (IPW) were used to reweight the outcomes data. The IPW was calculated as the ratio of two probabilities: the marginal probability of having a 2^nd^ CPP pregnancy as the numerator, and the conditional probability of a 2^nd^ CPP pregnancy as the denominator, where the conditional probability model included a wide range of variables as described in Supplementary Table 1. The Kaplan-Meier, and the log(-log(S(t)) plots were used to visually assess proportionality, followed by Schoenfeld residual analyses to confirm that the proportional hazards assumption was satisfied for both exposures.

*Analyses with primary exposure ICWC:* ICWC was categorized into quintiles, to identify any potential non-linear associations between the exposure and the outcome. The risks of all-cause mortality, associated with quintiles of ICWC were estimated using IPW weighted Cox proportional hazard (PH) model. Three covariate adjustment scenarios were applied: (1) unadjusted, (2) adjusted for confounders, including socio-demographic characteristics, smoking status, reproductive and medical history, and study site; and (3) additionally adjusted for interconception interval (ICI) i.e., the time difference between the LMP of the 1^st^ and 2^nd^ CPP pregnancies, to assess if time between two pregnancies contributed to weight change.

Multiple sensitivity analyses were conducted. First, to elucidate the effect of extreme weight loss or weight gain, we separated the top and bottom 1% of ICWC values following an initial review of the data distribution, thus rendering 7 different categories of ICWC. Second, we conducted a stratified analysis wherein the quintiles of ICWC were reassessed within BMI categories: <18.5, 18.5-24.9, 25.0-29.9, ≥30.0. However, given the small sample sizes within some of the BMI category-specific quintiles, the risks of all-cause mortality were estimated only for those with normal (18.5-24.9) pre-pregnancy BMI (70.5%, n=5754).

Cause-specific mortality risks due to cardiovascular, diabetes and kidney conditions associated with quintiles of ICWC were estimated using cause-specific hazard models, which model the instantaneous hazard of death from a specific cause among individuals who remain event-free, assuming independent censoring by competing causes conditional on covariates(37). Cause-specific mortality risks associated with the BMI-specific quintiles were not implemented due to small sizes. Details of the model specifications are provided in Supplementary Table 1.

*Analyses with secondary exposure IPWC:* IPWC was also categorized into quintiles. The risks of all-cause mortality, associated with quintiles of IPWC and BMI-specific quintiles of IPWC were estimated using similar modeling approaches as the ICWC analyses. The adjusted models were additionally accounted for 1^st^ pregnancy GWG and model 3 alternatively adjusted for the IPI i.e., the time difference between 1^st^ pregnancy delivery date and LMP of the 2^nd^ pregnancy.

We completed multiple sensitivity analyses on IPWC. Given that the majority of the study participants lost weight and only a small fraction had weight gain during IPI, a third scenario distinguishing weight gain as a separate category from the 5^th^ quintile of IPWC was also evaluated to tease out the risk associated with postpartum weight gain. This third scenario was also implemented to models with BMI-specific quintiles of IPWC. In addition, we repeated the above all-cause mortality analysis with IPWC quintiles, and with the weight gain group separated out from Q5, to include additional covariate adjustment and account for three potential confounding mechanisms. First, we adjusted for birth weight as a covariate given that our derivation of IPWC incorporates weight of the baby and therefore may account partially for the weight change patterns observed during IPI in this study. Secondly, there might have been an impact of shorter gestation among women delivering preterm on IPWC measures as well as on mortality—women delivering preterm were likely to have gained less gestational weight, and preterm delivery is known to be associated with an increased mortality by cardiovascular disease(38). Therefore, we also ran a separate model including gestational age at delivery as a covariate. Finally, although pregnancy outcome is presumed to influence IPWC predominantly via postpartum experiences and behaviors, a direct pathway cannot be completely dismissed. Thus, we fitted a third model that included pregnancy outcome as an additional covariate.

## Results

For both ICWC and IPWC, Kaplan–Meier and log[–log S(t)] plots (Supplementary Figure 1) showed parallel curves across quintiles, and Schoenfeld residuals (Supplementary Figure 2) showed no systematic deviation from zero over time i.e., no evidence of heterogeneity over follow-up, indicating no violation of the proportional hazards assumption.

### Risk of mortality associated with ICWC

The majority of women (∼56%, n=4141) had a higher weight at the beginning of their 2^nd^ pregnancy than their 1^st^ pregnancy (Table 1) and the median ICWC was 0.91 kg [IQR:-0.45, 3.63] (Supplementary Figure 3). The distributions of most participant characteristics were similar across quintiles. Some notable exceptions include the increase in the percentage of non-smokers as ICWC increased, with the highest percentage of non-smokers in the highest quintile of ICWC. The interval between conceptions for most quintiles of ICWC (1^st^, 3^rd^ and 4^th^ quintiles) averaged around two years (∼24 months) but was slightly shorter for the 2^nd^ quintile (∼22 months) and longer (∼27 months) for the 5^th^ quintile of ICWC. The median follow-up for this study sample was 54 years [inter-quartile range (IQR): 47-56] and 41.7% (n=3405) died during the study period (Table 2).

**Table 1:**
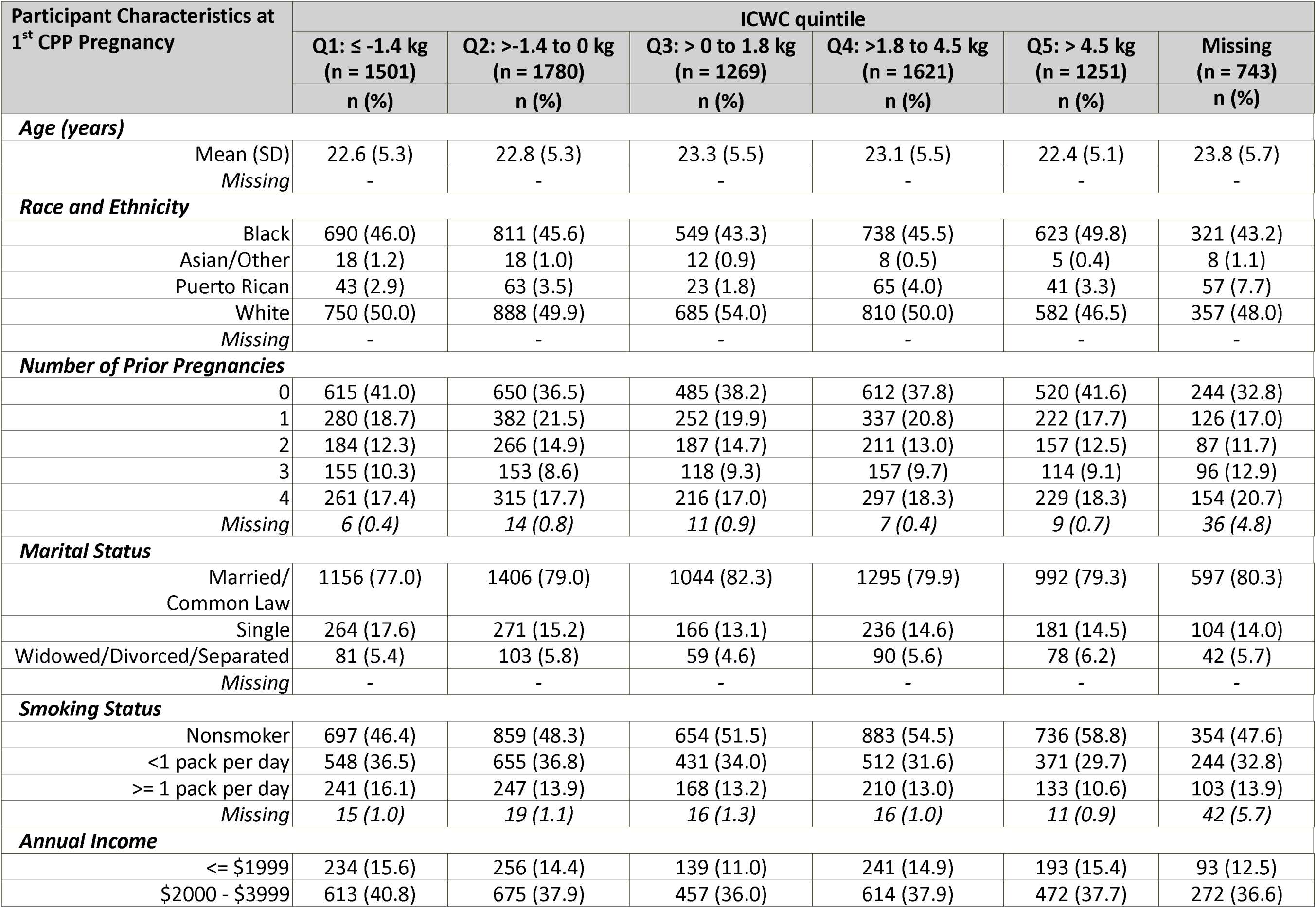
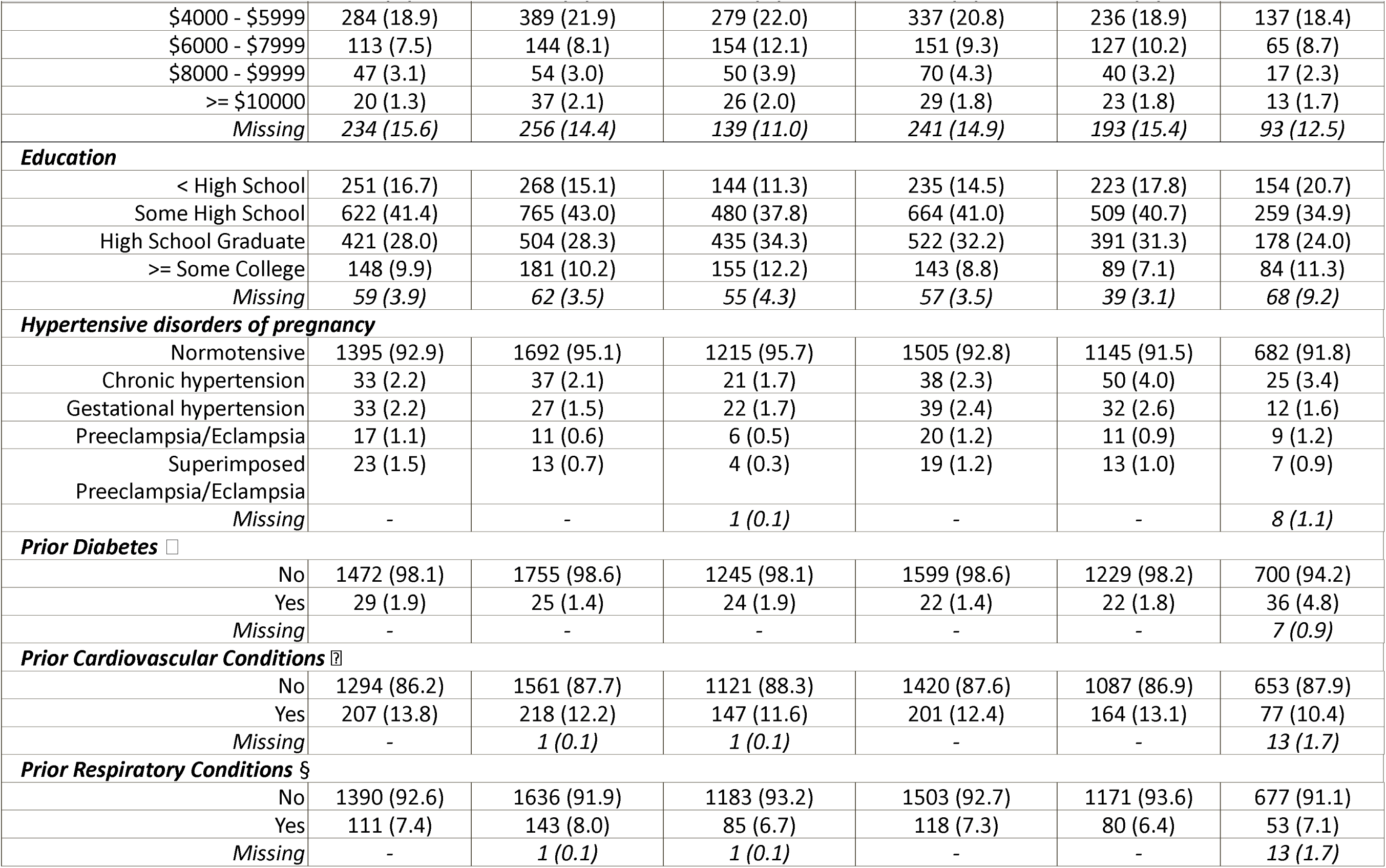
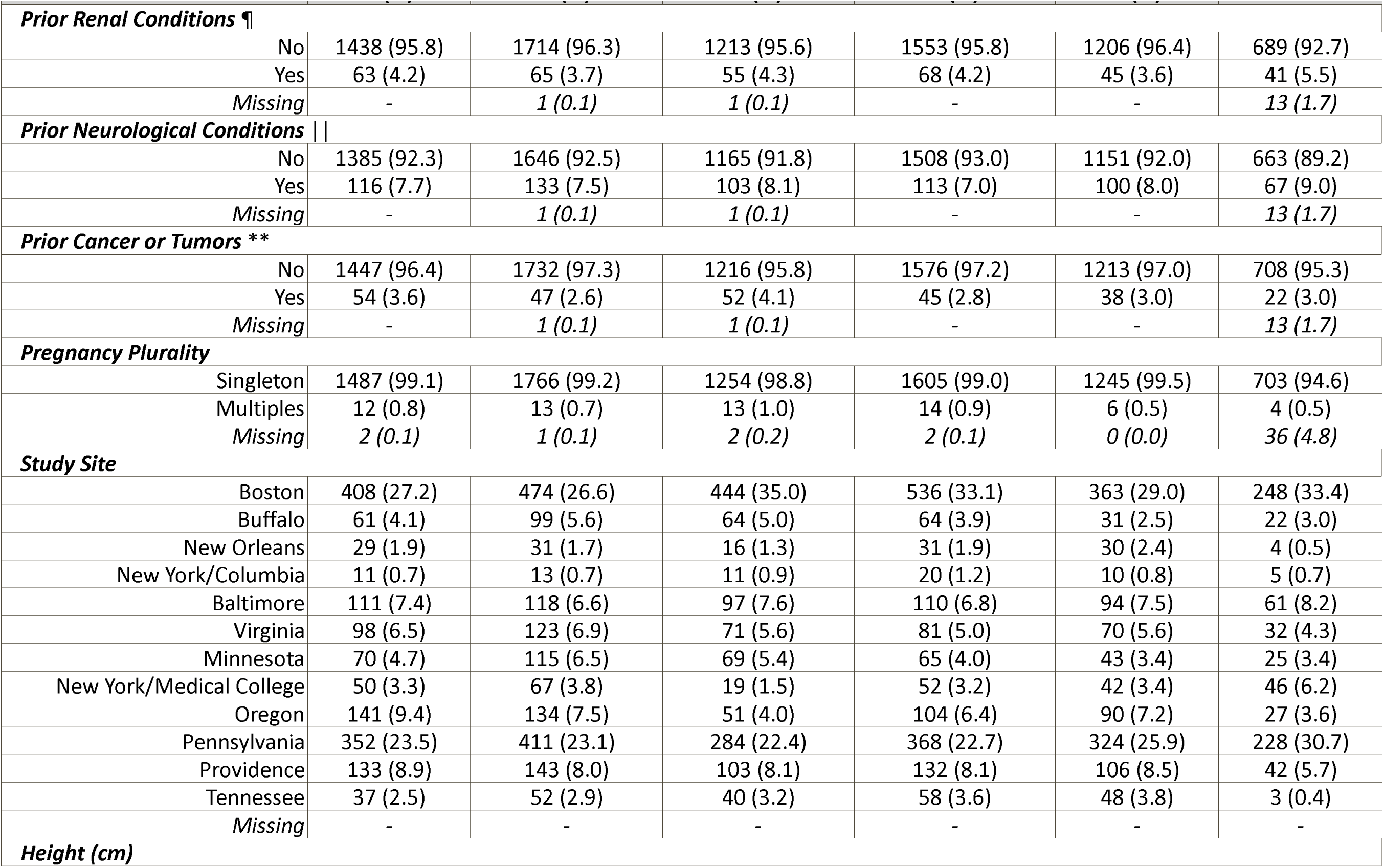
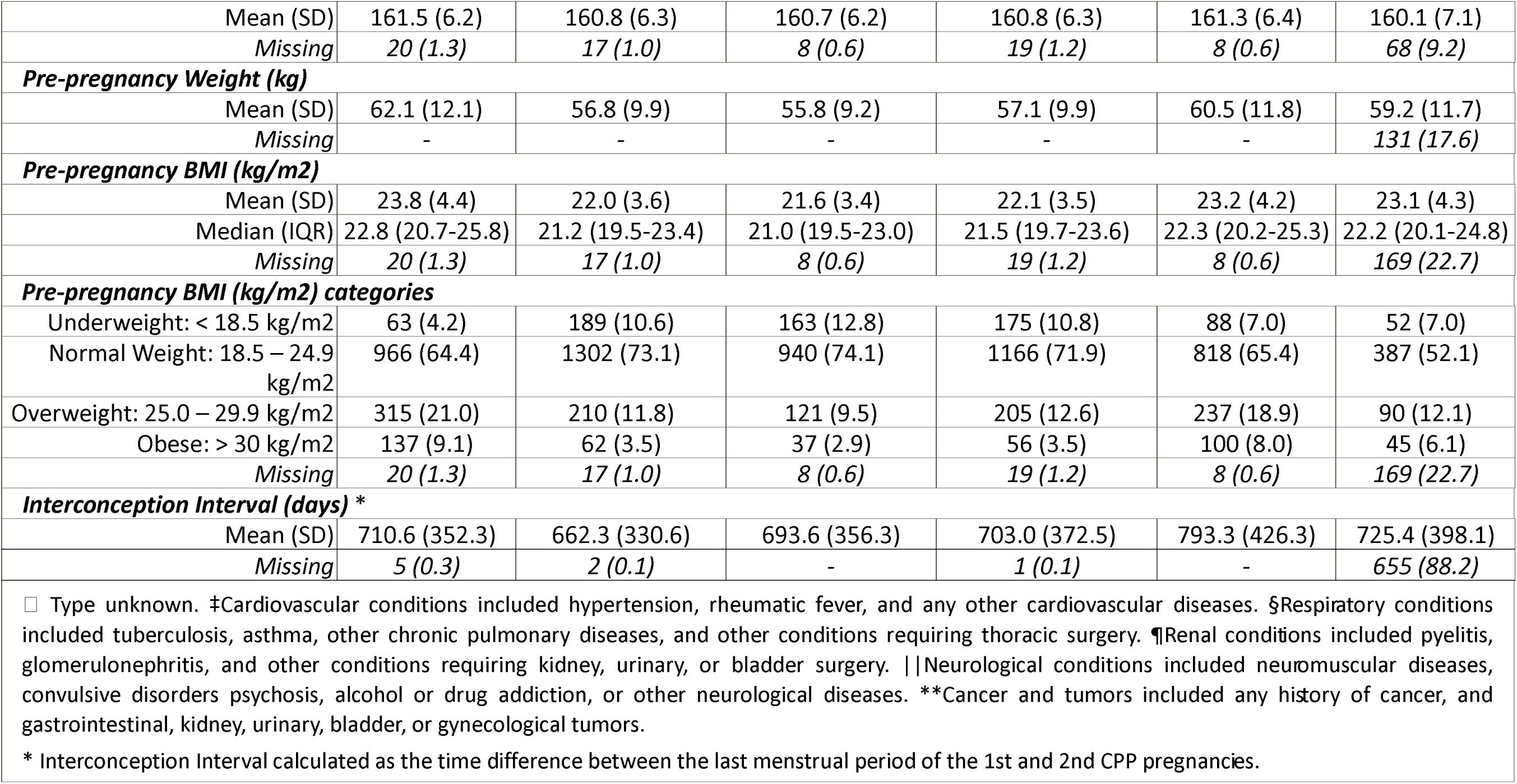
Summary of participant characteristics across quintiles of interconception weight change (ICWC)

**Table 2:** Distribution of outcome of interest across quintiles of interconception weight change (ICWC)

| Outcomes of Interest |  | ICWC quintile |  |  |  |  |  |
| --- | --- | --- | --- | --- | --- | --- | --- |
| | | Q1: $\leq -1.4$ kg<br>(n = 1501) | Q2: $>-1.4$ to 0 kg<br>(n = 1780) | Q3: $> 0$ to 1.8 kg<br>(n = 1269) | Q4: $>1.8$ to 4.5 kg<br>(n = 1621) | Q5: $> 4.5$ kg<br>(n = 1251) | Missing<br>(n = 743) |
|  |  | n (%) | n (%) | n (%) | n (%) | n (%) | n (%) |
| Vital Status | Alive | 865 (57.6) | 1067 (59.9) | 752 (59.3) | 941 (58.1) | 712 (56.9) | 423 (56.9) |
|  | Dead | 636 (42.4) | 713 (40.1) | 517 (40.7) | 680 (41.9) | 539 (43.1) | 320 (43.1) |
|  | Missing | - | - | - | - | - | - |
| Cause of Death | Cardiovascular | 162 (25.5) | 196 (27.5) | 119 (23.0) | 184 (27.1) | 149 (27.6) | 89 (27.8) |
|  | Diabetes | 28 (4.4) | 19 (2.7) | 22 (4.3) | 22 (3.2) | 27 (5.0) | 18 (5.6) |
|  | Kidney | 8 (1.3) | 8 (1.1) | 10 (1.9) | 14 (2.1) | 14 (2.6) | 5 (1.6) |
|  | Missing | 61 (4.1) | 68 (3.8) | 46 (3.6) | 64 (3.9) | 56 (4.5) | 30 (4.0) |

Results from the adjusted model (Figure 2) demonstrated that compared to ICWC > 0 kg to 1.8 kg (Q3), ICWC > −1.4 to 0 kg (Q2) was associated with lower all-cause mortality [aHR (95% CI): 0.85 (0.76 – 0.96)]. Adjusting for ICI did not yield any meaningful changes in the estimates.

**Figure 2:**
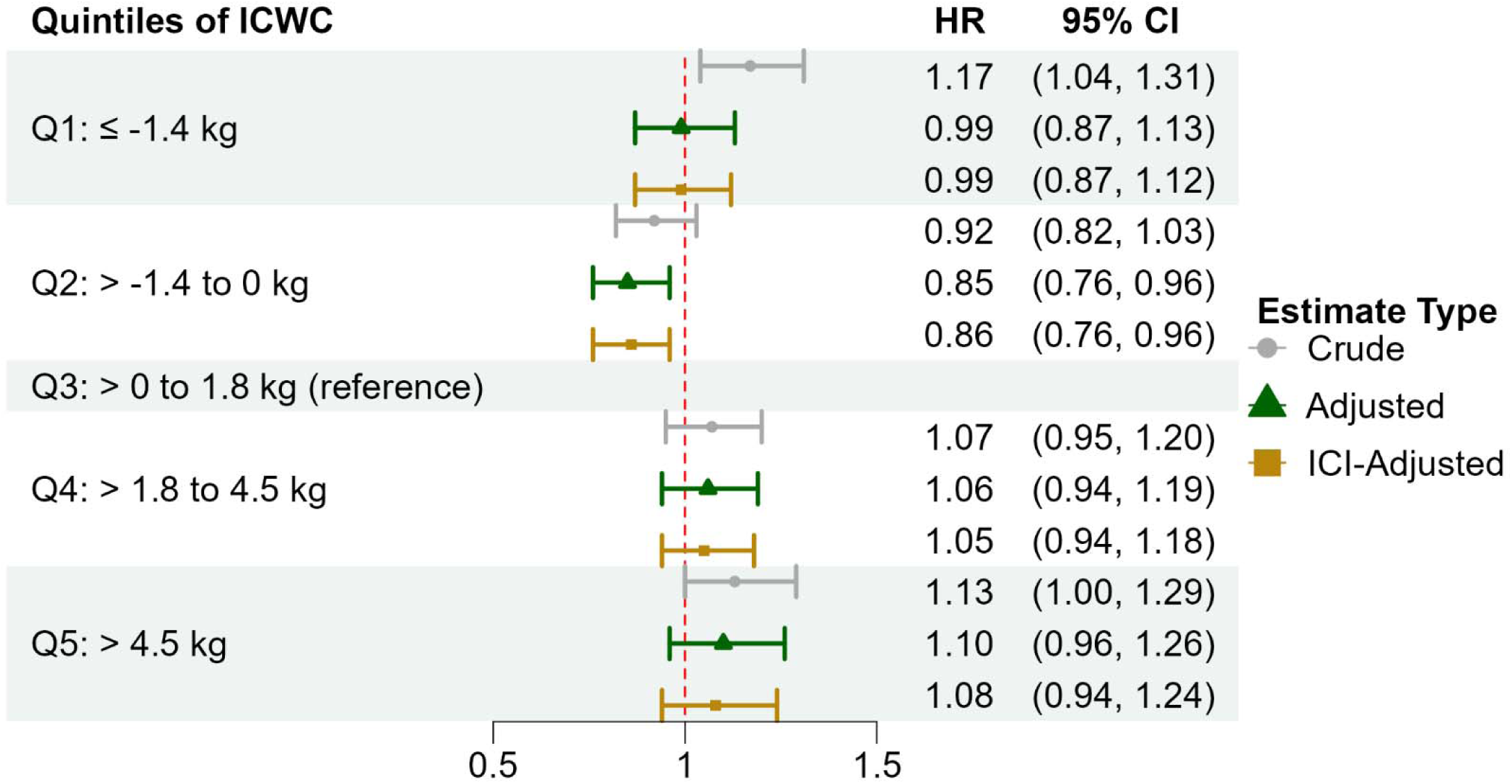
Risk of all cause mortality associated with quintiles of interconception weight change (ICWC) ICI: Interconception interval Adjusted for CPP 1^st^ Pregnancy variables: age, race and ethnicity, number of prior pregnancies, marital status, smoking status, annual income, education, prior diabetes, prior cardiovascular conditions, prior respiratory conditions, prior renal conditions, prior neurological conditions, prior cancer or tumors, pregnancy plurality, study site, and pre-pregnancy BMI

Distinguishing the top and bottom 1% of ICWC (Supplementary Figure 4) showed that very high ICWC > 17.2 kg [aHR (95% CI): 1.57 (1.10 – 2.23)] and very low ICWC ≤ −10.4 kg [aHR (95% CI): 1.15 (0.83 – 1.59)], were associated with higher all-cause mortality compared to Q3 of ICWC, albeit the results lacked precision.

Among those with normal pre-pregnancy BMI at their 1^st^ CPP pregnancy, ICWC > −0.9 and 0 kg was associated with a reduced risk of all-cause mortality [aHR (95% CI): 0.85 (0.73 – 0.99)] compared to ICWC > 0 to 1.8 kg (Supplementary Table 2).

Similar patterns of cause-specific mortality risk among women in Q2 and Q5 of ICWC were observed, although the estimates were imprecise due to limited sample sizes (Supplementary Table 3), with the notable exception of ICWC > −1.4 to 0 kg (Q2) which was associated with lower the risk of mortality due to diabetes [aHR (95% CI): 0.45 (0.23 – 0.87)].

### Risk of mortality associated with IPWC

Approximately 94% of participants experienced weight loss during the postpartum period (median IPWC: −8.16 kg [IQR:-4.9, −11.34]) (Supplementary Figure 3). Supplementary Table 4 describes the participant characteristics according to IPWC. Gestational weight gain was highest among those in Q1 (mean 13.8 kg [SD 4.5]) and lowest among those in Q5 (6.3 kg [4.8]). The IPI increased slightly from 1.2 years among those in Q1 to 1.4 years among those in Q5. We found no significant associations between IPWC and all-cause mortality (Figure 3). Nevertheless, reduced risks of all-cause mortality were suggested among those in Q1, Q2, and Q4 compared to Q3 of IPWC. Adjusting for IPI did not yield any significant changes in the estimates.

**Figure 3:**
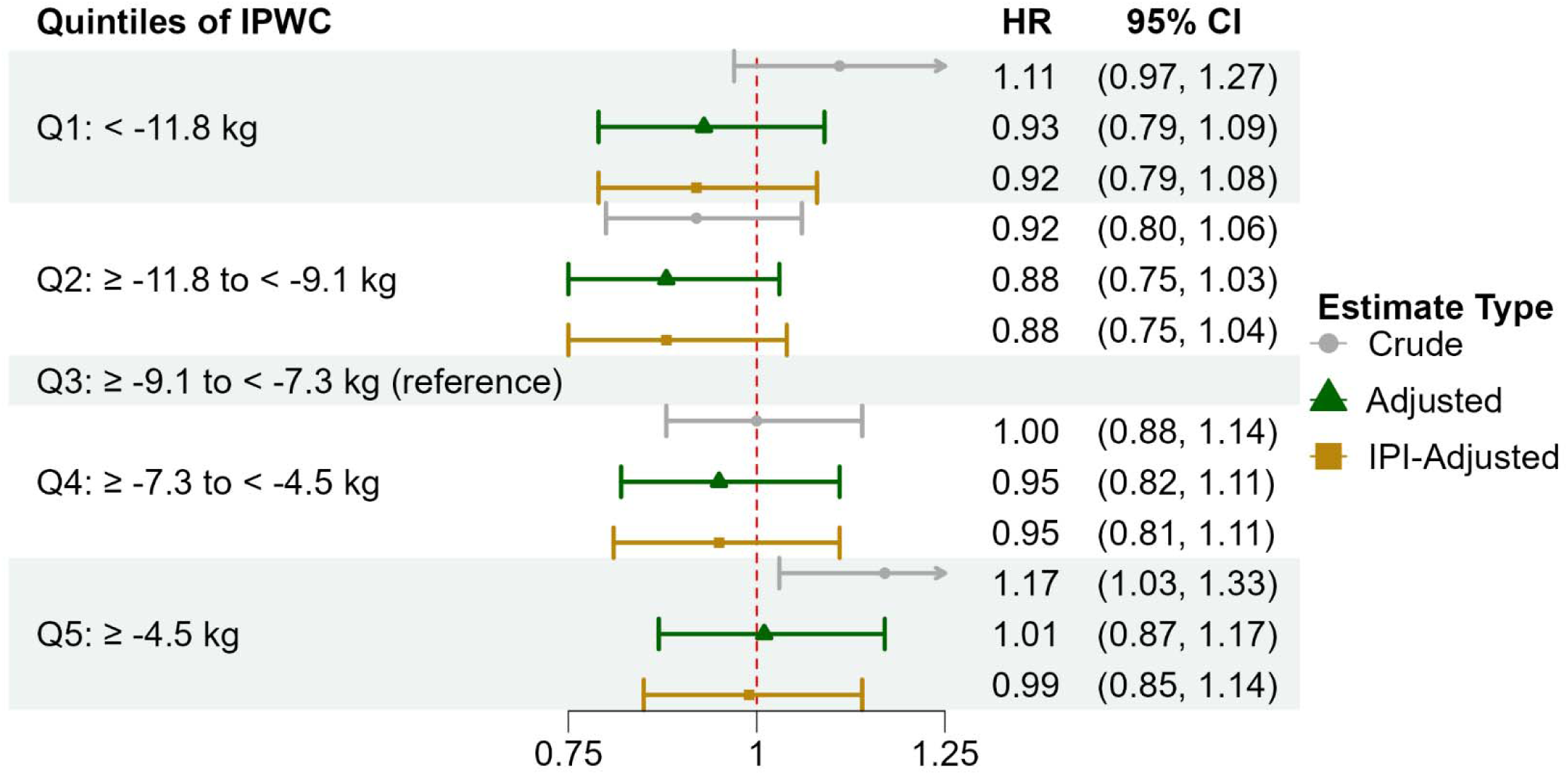
Risk of all cause mortality associated with quintiles of Interpregnancy Weight Change (IPWC) IPI: Interpregnancy interval Adjusted for CPP 1^st^ Pregnancy variables: age, race and ethnicity, number of prior pregnancies, marital status, smoking status, annual income, education, hypertensive disorders of pregnancy, prior diabetes, prior cardiovascular conditions, prior respiratory conditions, prior renal conditions, prior neurological conditions, prior cancer or tumors, pregnancy plurality, study site, pre-pregnancy BMI, and gestational weight gain

Further separation of the 5^th^ quintile into weight loss and gain (Supplementary Figure 5) showed an elevated but imprecise risk of all-cause mortality among those with weight gain [aHR (95% CI): 1.17 (0.95 – 1.43)]. Among those with normal pre-pregnancy BMI, weight loss of less than 5 kg during the IPI (i.e., Q5 of IPWC) was associated with a higher risk of all-cause mortality [aHR, 95% CI: 1.21, 1.02 – 1.43] (Supplementary Table 6). Additionally, those with weight retention or gain during the IPI (i.e. IPWC ≥ 0 kg) had an elevated risk of all-cause mortality [aHR, 95% CI: 1.35, 1.01 – 1.79]. Adjustment for birth weight, gestational age at delivery, and pregnancy outcome (Supplementary Table 7) yielded similar results.

The cause-specific mortality risks across IPWC quintiles appeared comparable; however, these results were unstable and imprecise due to the small number of events, precluding meaningful interpretation (Supplementary Table 8).

## Discussion

In a large, U.S. study with more than 50 years of follow-up after pregnancy, maternal postpartum weight retention independently was associated with long-term mortality risk, beyond those attributable to pre-pregnancy BMI and GWG. Specifically, we found that women who returned to their pre-pregnancy weight or maintain a weight slightly below that level, had a lower risk of all-cause mortality and experienced 55% lower diabetes-related mortality compared to those with moderate postpartum weight retention between pregnancies. Furthermore, women who entered a subsequent pregnancy at a higher weight than their pre-pregnancy baseline tended to have higher all-cause mortality, although estimates were not statistically significant. Our analysis of interpregnancy weight change, independent of gestational weight gain, suggested that participants who maintained or modestly reduced weight in the postpartum period had lower observed mortality, whereas those with weight retention or gain had higher observed mortality.

Our findings are novel and contribute to the growing body of literature on PPWR which highlights that women with high levels of PPWR face an increased risk of chronic conditions(12, 22–24). For instance, Kew et al. (2014) reported increased markers of adverse cardiometabolic health as early as 12 months postpartum, evidenced by higher mean adjusted diastolic blood pressure, insulin resistance, LDL cholesterol, and apolipoprotein B, along with declining adiponectin levels with higher postpartum weight gain trajectories(22). Studies with longer follow-up periods have reached similar conclusions regarding the long-term health risks associated with postpartum weight retention. For example, Soria-Contreras et al. (2020) found that increasing postpartum weight trajectories were associated with higher levels of central adiposity, inflammation, and LDL cholesterol levels in later postpartum years(12). Similarly, Kramer et al. (2024) reported that PPWR was associated not only with a higher risk of pre-diabetes/diabetes at 5-years postpartum, but also predicted worse characterization of cardiovascular risk indices such as triglycerides, high-/low-density lipoprotein, apolipoprotein-B, Matsuda index, insulin resistance, fasting glucose, and C-reactive protein(23). A study using data from the Danish National Birth Cohort (DNBC; 1997–2002) found elevated risks of hypertension and cardiovascular disease 16 years postpartum among women in the weight retention or gain groups(24). Due to the small sample size, which may have contributed to the lack of precision in our cause-specific results, we could not draw definitive conclusions about the increased risks of mortality from cardiovascular, and kidney conditions associated with ICWC or IPWC within this cohort. However, our finding of a 55% reduction in diabetes-related mortality from postpartum weight loss is notable. These results suggested that weight retention or gain during the postpartum period was associated with higher observed mortality, whereas weight loss was associated with lower observed mortality. Thus, our study not only complements previous studies that have linked PPWR to morbidity but also represents a novel contribution—being the first to map the association between PPWR and long-term mortality, and to suggest a potential survival benefit from healthy postpartum weight loss.

Our findings also underscore the need to implement postpartum weight management strategies across the full range of pre-pregnancy BMI categories. In 2021, the US Preventive Services Task Force found strong evidence(39–41) that behavioral interventions reduce the likelihood of excessive weight gain during pregnancy, recommending that healthcare providers offer behavioral counseling to pregnant women to promote healthy weight gain in line with the National Academy of Medicine (NAM) 2009 guidelines(42). However, such guidance for maintaining a healthy weight during the postpartum period remains limited. The few trials that have explored this area have focused on postpartum weight management for women living with overweight or obesity given their elevated risk of long-term health complications, as is evident in this systematic review(43) of 18 randomized controlled trials (2559 participants). However, BMI-specific analyses from this study suggest that women with low risk at baseline i.e., a normal pre-pregnancy BMI who had weight retention or gain during the postpartum period had higher observed mortality. Therefore, postpartum weight management interventions should also consider normal BMI groups and not just women with BMI > 24.9 kg/m^2^.

This study focused on pregnant women from the 1950s and 1960s, utilizing a historic cohort to examine the long-term health implications of PPWR, with follow-up extending over 50 years—an important and unique strength of this study given it is unfeasible to study long-term mortality utilizing contemporary cohorts. Yet, some characteristics, such as prevalence of smoking and underlying causes of mortality in CPP, which differ from current obstetric populations may limit generalizability. Other characteristics of the CPP cohort that may limit the generalizability of our findings include the underrepresentation of Hispanic women (2-3% in CPP compared to ∼26% in the 2023 U.S. obstetric population)(44), and a mean age of about 7 years younger than the current mean age of childbearing persons (∼30 years)(45). Contemporary obstetric populations also exhibit a higher prevalence of gestational weight gain (50%) exceeding recommendations(46–50) compared with the CPP cohort (11.6%). While these differences may hinder direct generalization of our findings, CPP’s rich covariate data and updated ICD-coded mortality enable future application of transportability methods(51) which can “transport,” results from the CPP-era obstetric population (source population) to the current obstetric population (target population), despite differences in key characteristics—a valuable direction for future research. We were also unable to evaluate the effect of gestational diabetes mellitus or impaired glucose tolerance during pregnancy, screening for which were limited during the study period(30, 52). Given the low prevalence of these conditions in our sample,(30) there was limited statistical power to assess them as independent covariates or effect modifiers. In addition to any residual confounding, self-reported pre-pregnancy weight may also contribute to bias, but this is likely minimal given the high correlation with measured pregnancy-related weights reported in other pregnancy cohorts(53, 54). Despite these limitations, the study’s analytical approach remains robust and is a key strength.

The analytical sample was restricted to women with at least two CPP-registered pregnancies; however, the use of inverse probability weighting (IPW) accounted for potential selection bias introduced by this restriction, allowing the results to remain generalizable to the overall CPP population. The study’s racial diversity, with nearly 50% of participants identifying as Black, is also a key strength, as much of the existing research on pregnancy exposures and long-term outcomes has predominantly focused on White populations. Finally, while the IPWC analyses were adjusted for IPI, the strength of the association between IPWC and long-term mortality may vary across different durations of IPI. Identifying such heterogeneity could help determine optimal time frames for postpartum weight management before a subsequent pregnancy. Future studies should explore potential effect modification by ICI, which was beyond the scope of this study due to sample size constraints. Similarly, while the IPWC analyses adjusted for GWG, the available sample size limits the ability to meaningfully evaluate GWG as an effect modifier, and this possibility should be explored in future studies.

## Conclusion

In conclusion, returning to pre-pregnancy weight or maintaining a weight slightly below that level, was associated with lower long-term mortality risk. This was true even among women with normal pre-pregnancy BMI. These findings underscore the benefit of postpartum weight loss and suggest the need for effective strategies that support healthy weight maintenance post-pregnancy. Future research with larger sample sizes and greater statistical power is needed to confirm these findings.

## Acknowledgments

Data from the Collaborative Perinatal Project is publicly available at https://www.archives.gov/research/electronic-records/nih.html (National Archives Identifier: 606622). Researchers interested in the linked mortality data should contact the NICHD (Edwina Yeung) for details on a data-sharing agreement and a confidentiality agreement with the National Death Index. The analysis codes are available from the corresponding author upon request.

