## Supplemental Materials for "Weight Change Between Pregnancies and Mortality Over 50 Years of Follow-up"

**Appendix A: Data Cleaning and Processing**

We preprocessed height and weight measurements by identifying and setting implausible values to missing based on predefined thresholds. Height discrepancies across pregnancies were resolved using the median value, while extreme values were flagged. Pre-pregnancy weight, delivery weight, and gestational weight gain (GWG) were set to missing if they fell outside biologically plausible ranges or deviated significantly (>±3 SD) from expected longitudinal trajectories. Weight measurements and rates of weight change between the two CPP pregnancies were further assessed in the context of both pregnancies, with extreme deviations flagged and set to missing. Using initial summary statistics of socio-demographic characteristics and other relevant information, we flagged and preprocessed height, pre-pregnancy weight, delivery weight, the 2^nd^ pregnancy starting weight and derived weight measurements data by setting them to missing based on the following strategies. First, for all CPP participants, we examined height across pregnancies. Height was discrepant by ≥ 0.0762 m (3 in) between at least two of the repeated height measurements for 2.9% (n=200) of participants with at least two measurements. Thus, for all participants with more than one recorded height, the median between all height measurements was utilized. The minimum height overall was also cleaned to set any values < 0.1^st^ percentile (1.2954 m; n=70) to missing; the 99.9^th^ percentile for height was plausible (1.8288 m), as was the upper limit (2.032 m) and therefore was left as is. Second, for any pregnancies with pre-pregnancy BMI <15 (n=80; 0.2%) or >50 (n=29; 0.1%) kg/m^2^ or pre-pregnancy weight <36.29 kg, the pre-pregnancy weight was set to missing. Weight at delivery and total gestational weight gain were also set to missing for any pregnancies with total gestational weight gain <-9.07 kg (-20 lbs.; n=123; 0.2%)) or < 36.29 kg (80 lbs.; n=20; <0.1%). Next, implausible pre-pregnancy weight (n=1118; 2.1%) and GWG (n=566; 1.1%) were assessed based on the context of their entire weight trajectory extracted from prenatal records using conditional percentiles^1^—implausible values were defined as those inconsistent with the expected weight distribution (>±3 SD) based on participants’ longitudinal prenatal trajectory. These records were also set to be missing. Lastly, all weight measurements were cleaned based on the context of both pregnancies’ weights and the time between the pregnancies. Weight measurements or their corresponding rate of weight change were set to missing for any records if either i) the difference or the rate of weight change between pre-pregnancy weights for 1^st^ and 2^nd^ pregnancies was < 0.1^th^ (-16.8 kg; n=8; -0.041 kg/week; n=7) or > 99.9^th^ (27.2 kg; n=9; 0.055 kg/week; n=8; <0.1%) percentile or ii) if the difference or rate of weight change between delivery weight for 1^st^ pregnancy and weight at the start of the 2^nd^ pregnancy was < 0.1^th^ (-28.1 kg; n=9; -3.175 kg/week; n=8; <0.1%) or > 99.9^th^ (14.1 kg; n=7; 0.558 kg/week; n=7; <0.1%).

1.	Yang S, Hutcheon JA. Identifying outliers and implausible values in growth trajectory data. *Ann Epidemiol*. 2016;26(1):77-80.e2. doi:10.1016/j.annepidem.2015.10.002

**Appendix B: Details on covariates used in different stages of the analysis in this study**

**Supplementary Table 1:** Specification of analytical models in terms of CPP 1^st^ pregnancy variables of interest

| **CPP 1^st^ Pregnancy Variables of Interest** | **Definition and Functional Form in Analytical Models** | **Analytical Model** | | |
| --- | --- | --- | --- | --- |
|  |  | **IPW ^1^** | **ICWC ^2^** | **IPWC ^3^** |
| Age (years) | Continuous | X | X | X |
| Race and Ethnicity | Categorical: White; Black; Puerto Rican; other ꝉ | X | X | X |
| Number of Prior Pregnancies | Categorical: 0; 1; 2; 3; 4 | X | X | X |
| Marital Status | Categorical: single; married/common law; other ꝉꝉ. | X | X | X |
| Smoking Status | Categorical: Non-smoker; <1 pack/day; ≥ 1 pack/day | X | X | X |
| Annual Income | Categorical: ≤$1999; $2000-3999; $4000-5999; $6000-7999; $8000-9999; ≥$10000. | X | X | X |
| Education | Categorical: <High School; Some high school; High School Graduate; Some College. | X | X | X |
| Hypertensive disorders of pregnancy | Categorical: Normotensive; Chronic hypertension; Gestational hypertension; Preeclampsia/ Eclampsia; Superimposed Preeclampsia/Eclampsia | X |  | X |
| Prior Diabetes | Binary: Yes; No | X | X | X |
| Prior Cardiovascular Conditions | Binary: Yes; No | X | X | X |
| Prior Respiratory Conditions | Binary: Yes; No | X | X | X |
| Prior Renal Conditions | Binary: Yes; No | X | X | X |
| Prior Neurologic Conditions | Binary: Yes; No | X | X | X |
| Prior Cancer or Tumors | Binary: Yes; No | X | X | X |
| Pregnancy Plurality | Binary: Singleton; Multiples | X | X | X |
| Study Site | Categorical: Boston (reference); Buffalo; New Orleans; New York/Columbia; Baltimore; Virginia; Minnesota; New York/Medical College; Oregon; Pennsylvania; Providence; Tennessee | X | X | X |
| Pre-pregnancy BMI (kg/m^2^) * | Continuous: calculated from self-reported pre-pregnancy weight and height. | X | X | X |
| Birth Weight (g) | Continuous: measured at delivery |  |  | X |
| Gestational Weight Gain (kg) | Continuous: calculated as the difference between weight measured at admission for delivery and self-reported pre-pregnancy weight |  |  | X |
| Gestational Age at Delivery (weeks) | Continuous: calculated based on date of delivery and date of LMP | X |  | X |
| Pregnancy outcome (denoting offspring(s) death/survival) ^ǂ^ | Categorical: 0 = Live birth, stillbirth (reference); 1 = Pregnancy loss; 2 = Neonatal death; 3 = Infant or child death | X |  | X |
| Year of registration into CPP | Continuous | X |  |  |
| Interconception Interval (in days) | Continuous: calculated as the time difference between the LMP of the 1st and 2nd CPP pregnancies |  | X^a^ |  |
| Interpregnancy Interval (in years) | Continuous: calculated as the time difference between 1st pregnancy delivery date and LMP of the 2nd pregnancy |  |  | X^b^ |
| ^1^ Inverse Probability Weighting Model; ^2^ Model with ICWC as exposure; ^3^ Model with IPWC as exposure  ꝉ Race and ethnicity were self-reported by participants during study enrollment through in-person interviews. The original CPP data collection forms categorized race as ‘White,’ ‘Negro’ (herein referred to as ‘Black’), ‘Oriental’ (herein referred to as ‘Asian’), ‘Puerto Rican,’ and ‘Other.’ In this manuscript, the ‘Asian’ and ‘Other’ categories were combined into a single ‘Other’ group due to the small number of participants who identified as Asian.  ꝉꝉ Other=widowed, divorced, separated  * BMI-stratified sensitivity analyses models exclude Pre-pregnancy BMI as a covariate. Instead, these models were only run among the Normal pre-pregnancy BMI group.  ^ǂ^ Outcome codes were coded in the original CPP data collection forms as, 00 = Liveborn, still living; 01 = Abortion (≤19 weeks gestation); 02 = Abortion; 03 = Mole; 11 = Stillbirth (≥20 weeks gestation); 12 = Stillbirth (PRB Review); 29 = Fetal death, type unknown; 30 = Fetal death under 24 hours of age; 31 - 37 = Fetal death between day 1-7; 38 = Fetal death between 8–27 days; 39 = Neonatal death, time unknown; 40 = 28 days through 1 year; 51 = 1–2 years; 52 = 2 years 1 day to 3 years; 53 = 3 years 1 day to 4 years; 54 = 4 years 1 day to 5 years; 55 = 5 years 1 day to 6 years; 56 = 6 years 1 day to 7 years; 57 = 7 years 1 day to 8 years; 58 = Over 8 years; 59 = Child death, time unknown. In this manuscript, 00 = Liveborn, still living was coded as ‘0’ and is the reference group; outcome codes 01, 02, 03, 11, 12, 29 were collapsed into the group ‘1’ denoting the Pregnancy loss group; outcome codes 30, 31 – 37, 38, 39 were collapsed into the group ‘2’ denoting the Neonatal death group; outcome codes 40, 51, 52, 53, 54, 55, 56, 57, 58, 59 were collapsed into the group ‘3’ denoting the Infant or child death group.  ^a^ Included only for the ICWC model that was adjusted for ICI  ^b^ Included only for the IPWC model that was adjusted for IPI | | | | |

**Appendix C: Preliminary Analyses of Exposures and Outcomes**

**Supplementary Figure 1:** Visual Assessment of Proportionality Assumption with respect to ICWC and IPWC Quintiles (Imputed dataset 1)

**Time to Death by ICWC quintiles**

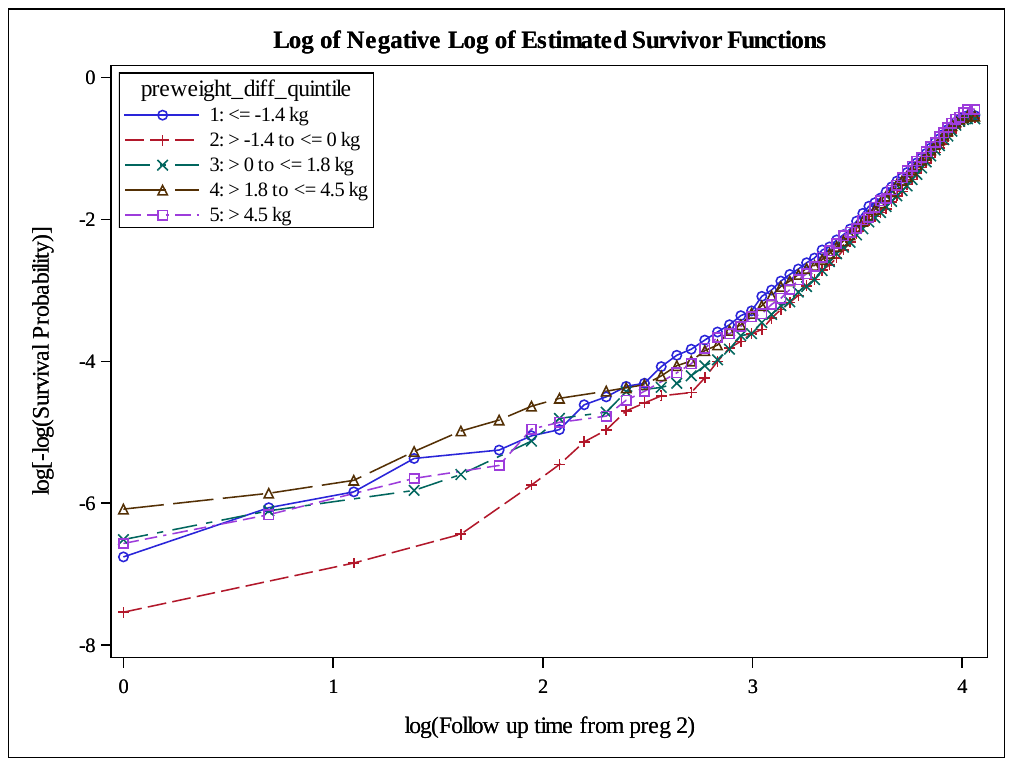

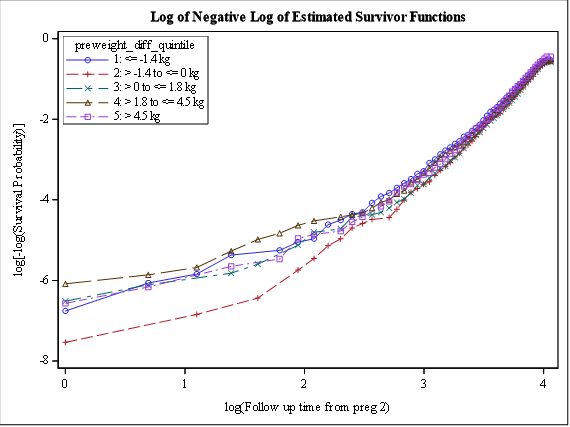

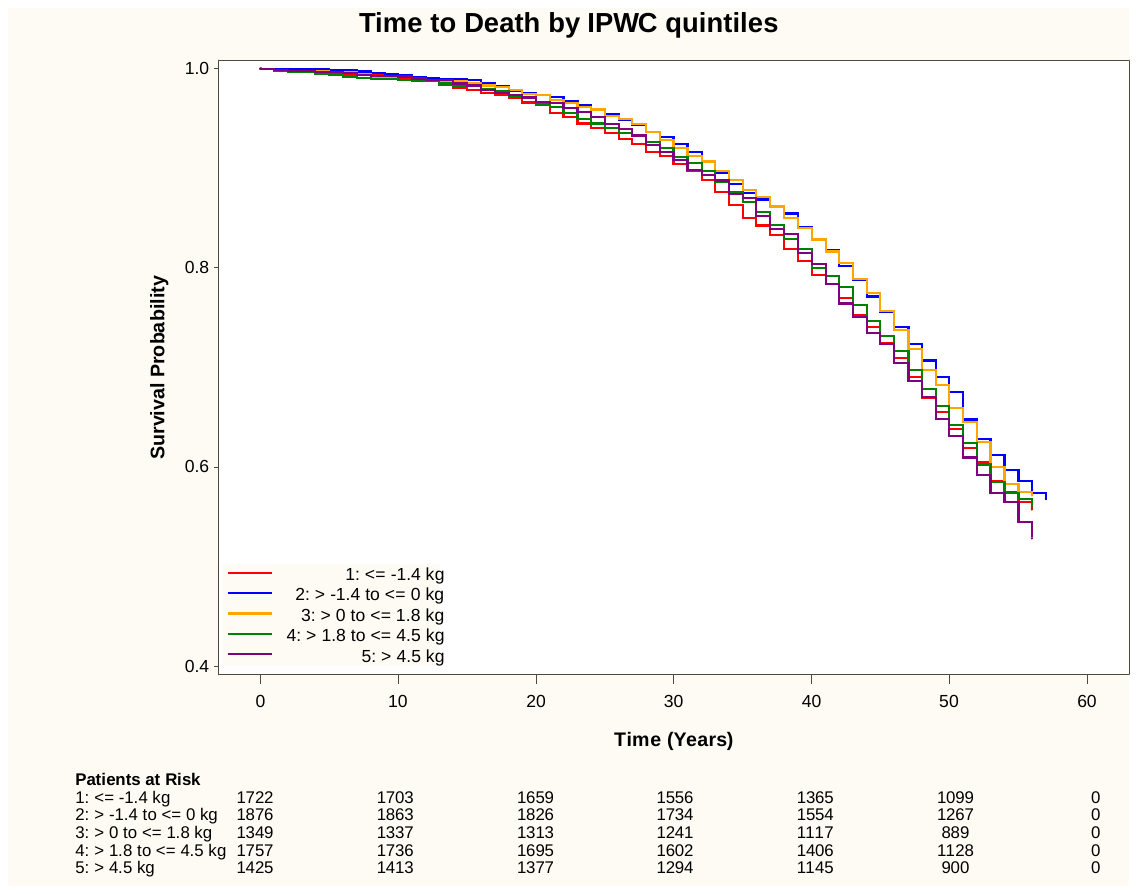

**ICWC quintiles**

**Time to Death by IPWC quintiles**

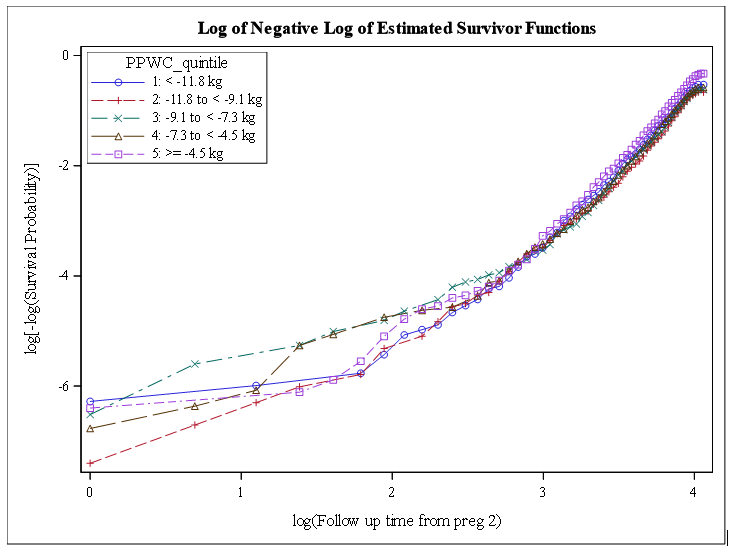

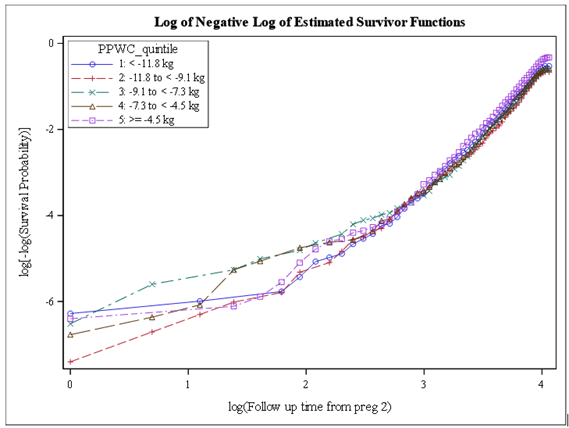

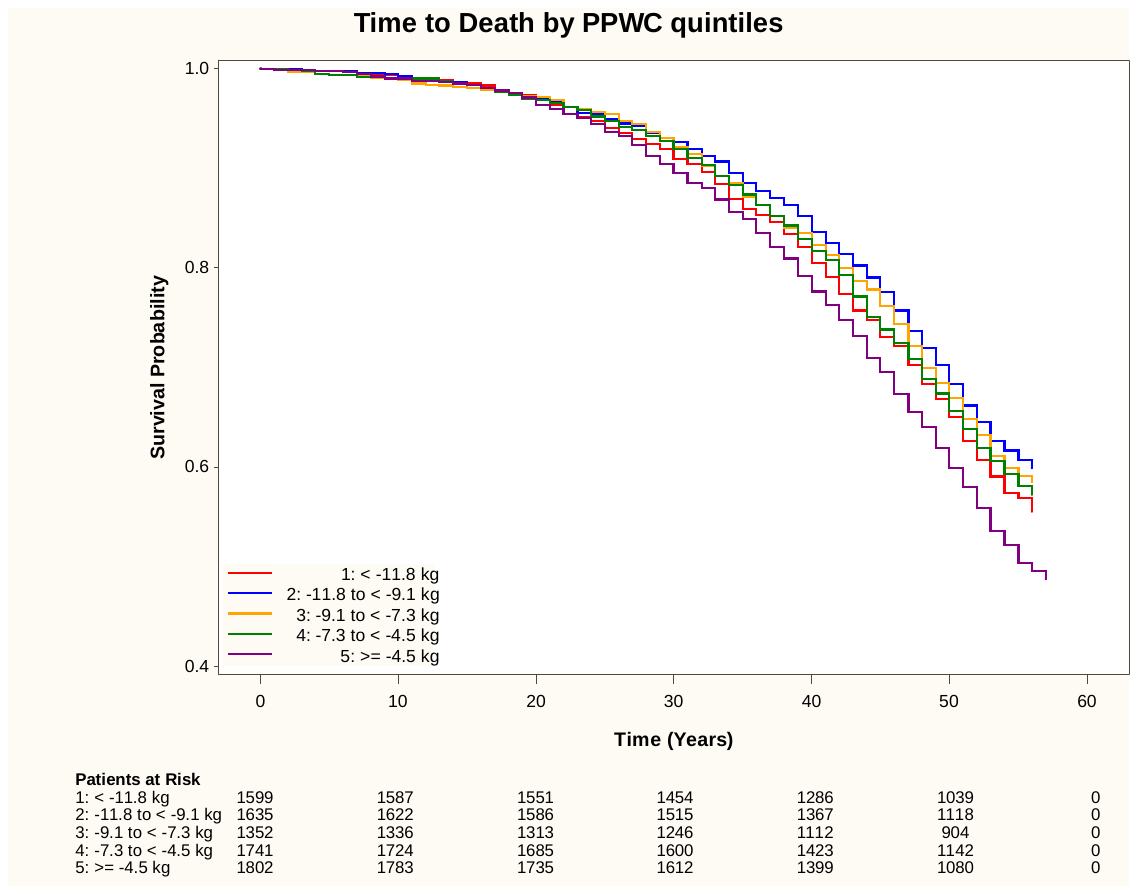

**IPWC quintiles**

**Supplementary Figure 2:** Schoenfeld residual plots with respect to ICWC and IPWC Quintiles (Imputed dataset 1)

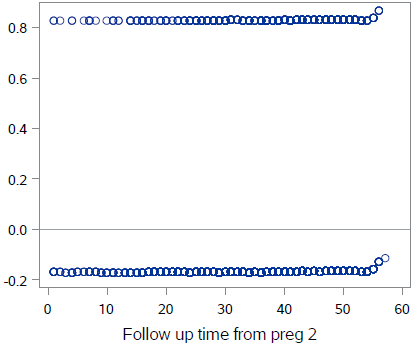

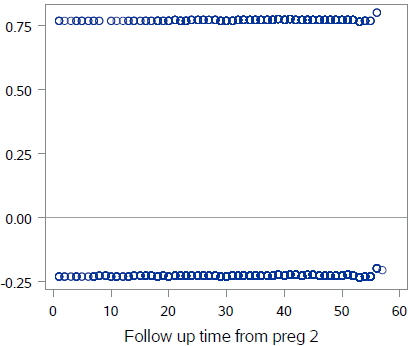

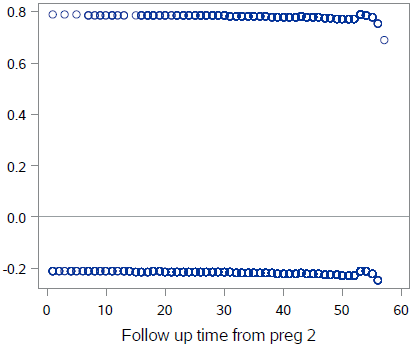

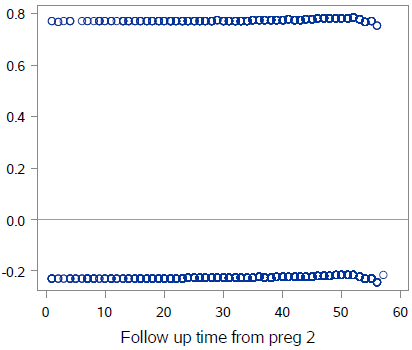

**ICWC Quintile 5 vs Quintile 3**

**ICWC Quintile 4 vs Quintile 3**

**ICWC Quintile 2 vs Quintile 3**

**ICWC Quintile 1 vs Quintile 3**

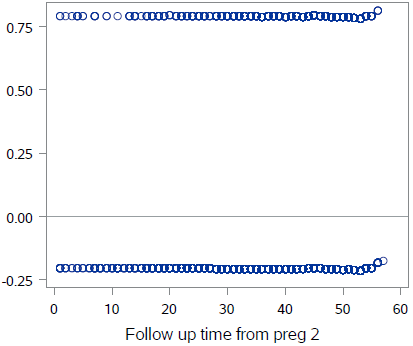

**IPWC Quintile 2 vs Quintile 3**

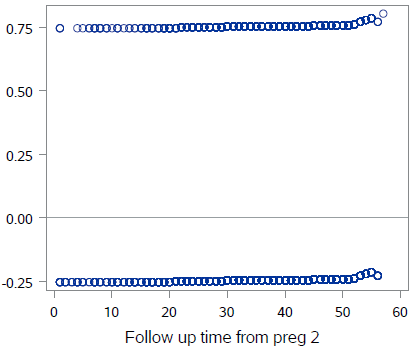

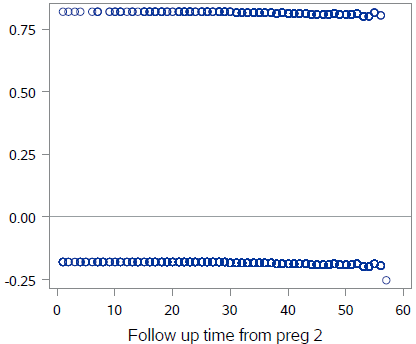

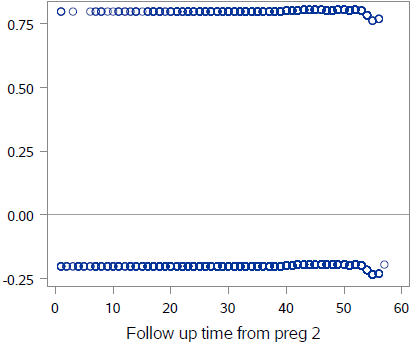

**IPWC Quintile 5 vs Quintile 3**

**IPWC Quintile 4 vs Quintile 3**

**IPWC Quintile 1 vs Quintile 3**

**Supplementary Figure 3:** Distribution of interconception weight change (ICWC) and interpregnancy weight change (IPWC)

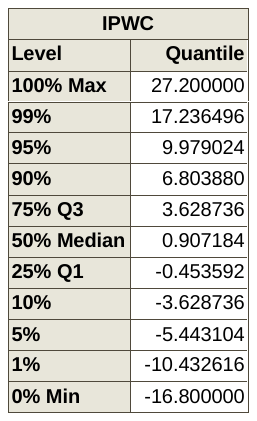

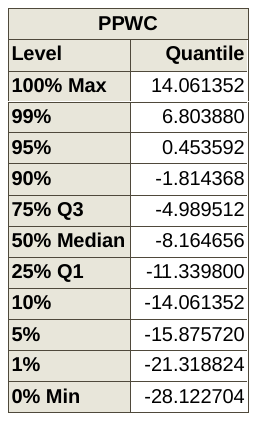

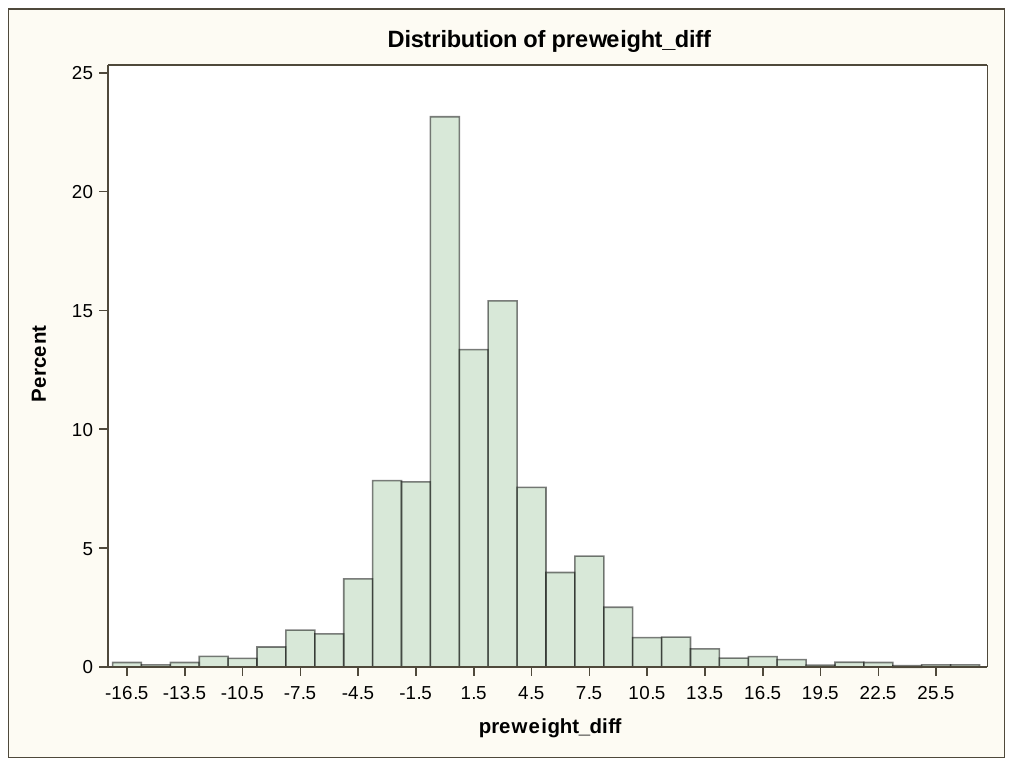

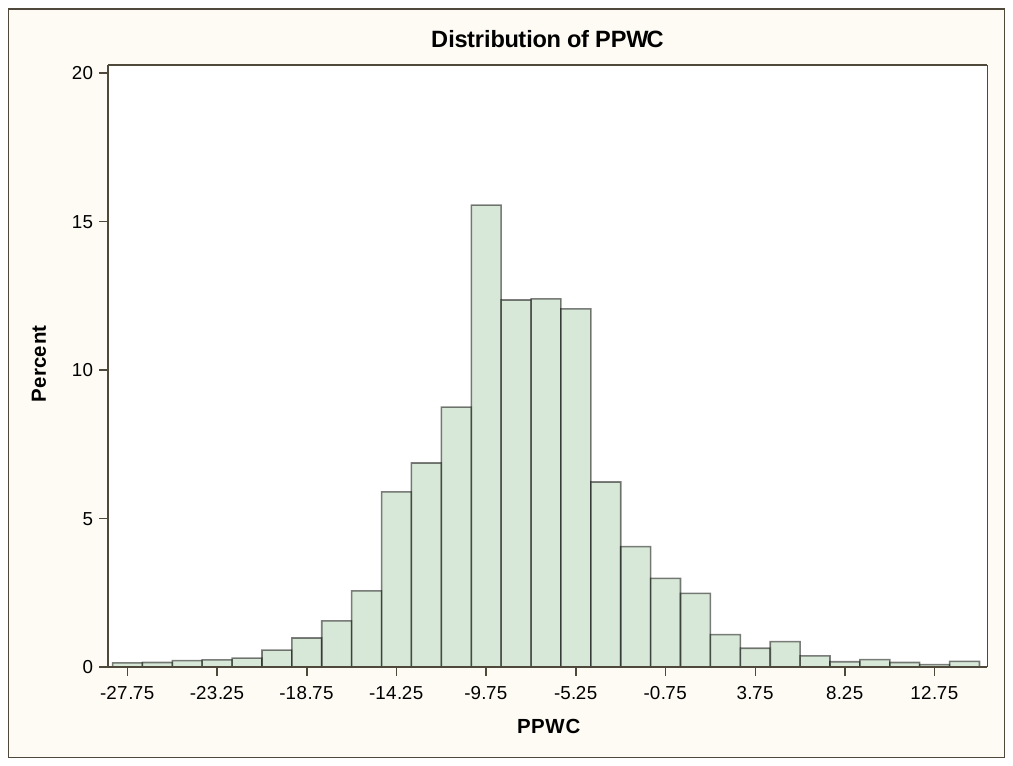

**IPWC**

**ICWC**

**Appendix D: Findings from sensitivity and cause-specific mortality analyses associated with interconception weight change (ICWC)**

**Supplementary Figure 4:** Risk of all cause mortality associated with quintiles of interconception weight change (ICWC) where the top and bottom 1% were further separated out

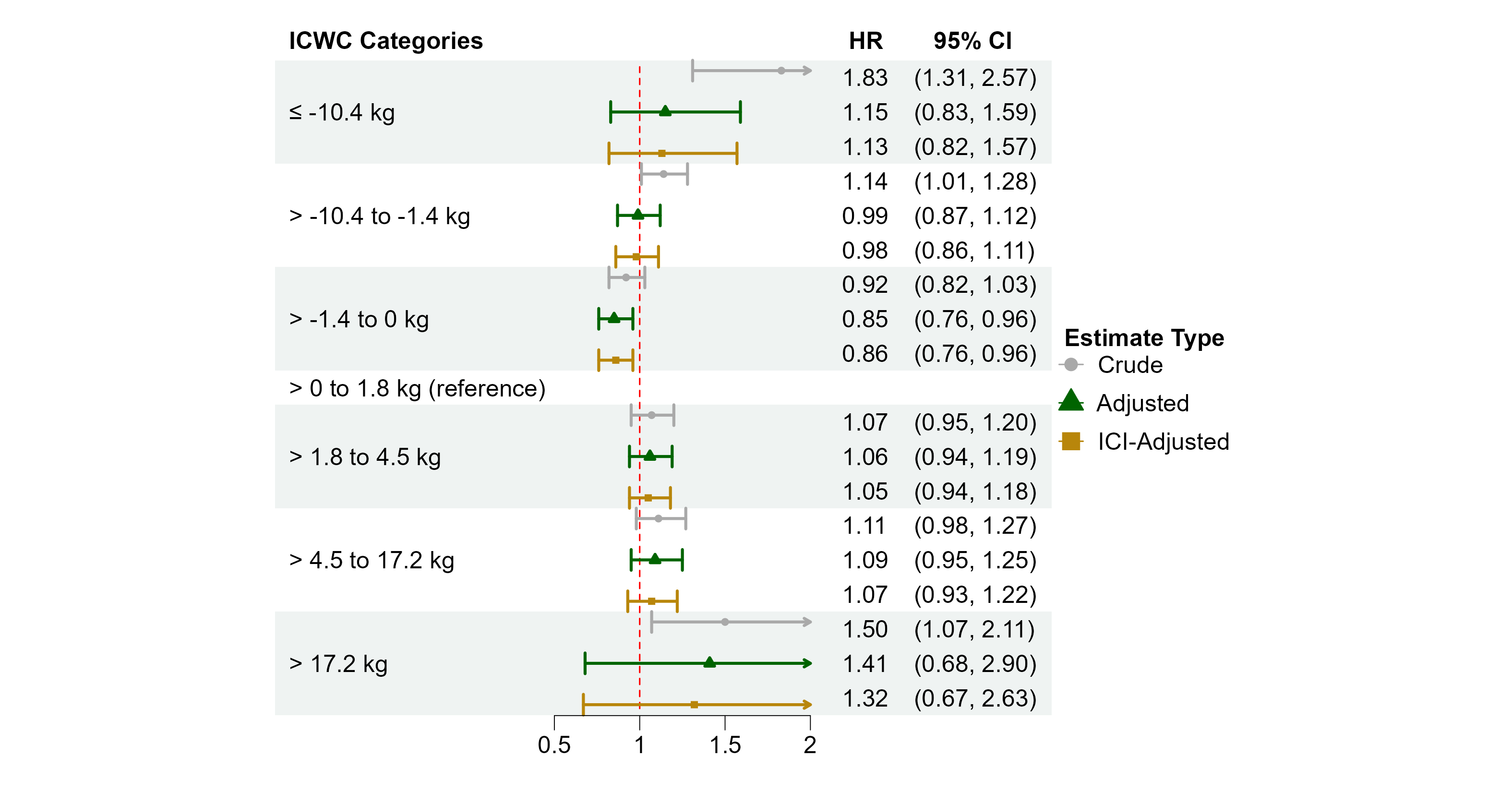

ICI: Interconception interval

Adjusted for CPP 1st Pregnancy variables: age, race and ethnicity, number of prior pregnancies, marital status, smoking status, annual income, education, prior diabetes, prior cardiovascular conditions, prior respiratory conditions, prior renal conditions, prior neurological conditions, prior cancer or tumors, pregnancy plurality, study site, and pre-pregnancy BMI

**Supplementary Table 2:** All-cause mortality risks associated with BMI-specific quintiles of interconception weight change (ICWC) for those with Normal pre-pregnancy BMI

| **Exposure** | **Category**  **(in kg)** | **Unadjusted** | **Adjusted *** | |
| --- | --- | --- | --- | --- |
|  |  | **HR (95% CI)** | **HR (95% CI) adjusted without ICI** | **HR (95% CI) adjusted with ICI** |
| **BMI-specific quintiles of ICWC** | Q1: ≤ -0.9 | 1.04 (0.90-1.20) | 1.03 (0.87-1.22) | 1.01 (0.85-1.20) |
|  | Q2: > -0.9 to 0 | 0.87 (0.76-1.01) | 0.85 (0.73-0.99) | 0.86 (0.74-1.00) |
|  | Q3: > 0 to 1.8 | reference | | |
|  | Q4: > 1.8 to 4.5 | 1.07 (0.94-1.23) | 1.08 (0.94-1.25) | 1.07 (0.93-1.23) |
|  | Q5: > 4.5 | 1.13 (0.95-1.33) | 1.17 (0.99-1.40) | 1.14 (0.96-1.36) |
| * Adjusted for CPP 1^st^ Pregnancy variables: age, race and ethnicity, number of prior pregnancies, marital status, smoking status, annual income, education, prior diabetes, prior cardiovascular conditions, prior respiratory conditions, prior renal conditions, prior neurological conditions, prior cancer or tumors, pregnancy plurality, and study site | | | | |

**Supplementary Table 3:** Cause-specific mortality associated with quintiles of interconception weight change (ICWC)

| **Cause of Death** | **Exposure**  **(quintiles of ICWC in kg)** | **Unadjusted** | **Adjusted *** | |
| --- | --- | --- | --- | --- |
|  |  | **HR (95% CI)** | **HR (95% CI) adjusted without ICI** | **HR (95% CI) adjusted with ICI** |
| **Cardiovascular** | Q1: ≤ -1.4 | 1.23 (0.98-1.54) | 1.00 (0.79-1.27) | 1.00 (0.79-1.26) |
|  | Q2: > -1.4 to 0 | 0.98 (0.77-1.24) | 0.88 (0.69-1.13) | 0.89 (0.69-1.14) |
|  | Q3: > 0 to 1.8 | reference | | |
|  | Q4: > 1.8 to 4.5 | 1.08 (0.86-1.36) | 1.03 (0.81-1.29) | 1.02 (0.81-1.29) |
|  | Q5: > 4.5 | 1.12 (0.86-1.46) | 1.04 (0.80-1.34) | 1.01 (0.79-1.30) |
| **Diabetes** | Q1: ≤ -1.4 | 1.22 (0.71-2.10) | 0.76 (0.43-1.34) | 0.75 (0.42-1.33) |
|  | Q2: > -1.4 to 0 | 0.59 (0.33-1.06) | 0.45 (0.23-0.87) | 0.45 (0.24-0.87) |
|  | Q3: > 0 to 1.8 | reference | | |
|  | Q4: > 1.8 to 4.5 | 1.17 (0.67-2.04) | 0.94 (0.51-1.74) | 0.93 (0.51-1.72) |
|  | Q5: > 4.5 | 1.80 (1.06-3.09) | 1.42 (0.82-2.47) | 1.39 (0.79-2.43) |
| **Kidney** | Q1: ≤ -1.4 | 0.58 (0.17-2.02) | 0.41 (0.13-1.32) | 0.41 (0.13-1.31) |
|  | Q2: > -1.4 to 0 | 0.76 (0.28-2.04) | 0.68 (0.26-1.81) | 0.68 (0.26-1.81) |
|  | Q3: > 0 to 1.8 | reference | | |
|  | Q4: > 1.8 to 4.5 | 0.94 (0.41-2.17) | 1.00 (0.41-2.45) | 0.99 (0.41-2.42) |
|  | Q5: > 4.5 | 1.47 (0.67-3.26) | 1.20 (0.51-2.80) | 1.18 (0.51-2.75) |
| * Adjusted for CPP 1^st^ Pregnancy variables: age, race and ethnicity, number of prior pregnancies, marital status, smoking status, annual income, education, prior diabetes, prior cardiovascular conditions, prior respiratory conditions, prior renal conditions, prior neurological conditions, prior cancer or tumors, pregnancy plurality, study site, and pre-pregnancy BMI | | | | |

**Appendix E: Findings related to the secondary exposure variable**

**Supplementary Table 4:** Summary of participant characteristics across quintiles of interpregnancy weight change (IPWC)

| **Participant Characteristics at 1^st^ CPP Pregnancy** | **IPWC quintile** | | | | | |
| --- | --- | --- | --- | --- | --- | --- |
|  | **Q1: < -11.8 kg (n = 1329)** | **Q2: ≥-11.8 to <-9.1 kg (n = 1436)** | **Q3: ≥-9.1 to <-7.3 kg (n = 1218)** | **Q4: ≥-7.3 to <-4.5 kg (n = 1553)** | **Q5: ≥-4.5 kg (n = 1455)** | **Missing (n = 1174)** |
|  | **n (%)** | **n (%)** | **n (%)** | **n (%)** | **n (%)** | **n (%)** |
| ***Age (years)*** | | | | | | |
| Mean (SD) | 22.3 (5.0) | 22.3 (5.1) | 22.9 (5.2) | 23.0 (5.5) | 23.6 (5.6) | 23.7 (5.7) |
| Missing | - | - | - | - | - | - |
| ***Race and Ethnicity*** | | | | | | |
| Black | 598 (45.0) | 608 (42.3) | 513 (42.1) | 697 (44.9) | 757 (52.0) | 559 (47.6) |
| Asian/Other | 15 (1.1) | 14 (1.0) | 15 (1.2) | 8 (0.5) | 6 (0.4) | 11 (0.9) |
| Puerto Rican | 37 (2.8) | 37 (2.6) | 46 (3.8) | 43 (2.8) | 52 (3.6) | 77 (6.6) |
| White | 679 (51.1) | 777 (54.1) | 644 (52.9) | 805 (51.8) | 640 (44.0) | 527 (44.9) |
| *Missing* | *-* | *-* | *-* | *-* | *-* | *-* |
| ***Number of Prior Pregnancies*** | | | | | | |
| 0 | 553 (41.6) | 633 (44.1) | 514 (42.2) | 586 (37.7) | 482 (33.1) | 358 (30.5) |
| 1 | 256 (19.3) | 291 (20.3) | 232 (19.0) | 323 (20.8) | 289 (19.9) | 208 (17.7) |
| 2 | 155 (11.7) | 179 (12.5) | 168 (13.8) | 225 (14.5) | 209 (14.4) | 156 (13.3) |
| 3 | 131 (9.9) | 119 (8.3) | 105 (8.6) | 140 (9.0) | 154 (10.6) | 144 (12.3) |
| 4 | 230 (17.3) | 210 (14.6) | 194 (15.9) | 274 (17.6) | 298 (20.5) | 266 (22.7) |
| *Missing* | *4 (0.3)* | *4 (0.3)* | *5 (0.4)* | *5 (0.3)* | *23 (1.6)* | *42 (3.6)* |
| ***Marital Status*** | | | | | | |
| Married/Common Law | 1018 (76.6) | 1129 (78.6) | 1006 (82.6) | 1236 (79.6) | 1159 (79.7) | 942 (80.2) |
| Single | 221 (16.6) | 226 (15.7) | 160 (13.1) | 236 (15.2) | 212 (14.6) | 167 (14.2) |
| Widowed/Divorced/Separated | 90 (6.8) | 81 (5.6) | 52 (4.3) | 81 (5.2) | 84 (5.8) | 65 (5.5) |
| *Missing* | *-* | *-* | *-* | *-* | *-* | *-* |
| ***Smoking Status*** | | | | | | |
| Nonsmoker | 631 (47.5) | 728 (50.7) | 653 (53.6) | 803 (51.7) | 811 (55.7) | 557 (47.4) |
| <1 pack per day | 521 (39.2) | 518 (36.1) | 386 (31.7) | 519 (33.4) | 433 (29.8) | 384 (32.7) |
| >= 1 pack per day | 172 (12.9) | 185 (12.9) | 169 (13.9) | 224 (14.4) | 182 (12.5) | 170 (14.5) |
| *Missing* | *5 (0.4)* | *5 (0.3)* | *10 (0.8)* | *7 (0.5)* | *29 (2.0)* | *63 (5.4)* |
| ***Annual Income*** | | | | | | |
| <= $1999 | 201 (15.1) | 215 (15.0) | 141 (11.6) | 209 (13.5) | 225 (15.5) | 165 (14.1) |
| $2000 - $3999 | 544 (40.9) | 545 (38.0) | 463 (38.0) | 570 (36.7) | 550 (37.8) | 431 (36.7) |
| $4000 - $5999 | 252 (19.0) | 290 (20.2) | 257 (21.1) | 351 (22.6) | 287 (19.7) | 225 (19.2) |
| $6000 - $7999 | 110 (8.3) | 144 (10.0) | 137 (11.2) | 137 (8.8) | 142 (9.8) | 84 (7.2) |
| $8000 - $9999 | 39 (2.9) | 54 (3.8) | 53 (4.4) | 57 (3.7) | 50 (3.4) | 25 (2.1) |
| >= $10000 | 19 (1.4) | 22 (1.5) | 37 (3.0) | 30 (1.9) | 26 (1.8) | 14 (1.2) |
| *Missing* | *164 (12.3)* | *166 (11.6)* | *130 (10.7)* | *199 (12.8)* | *175 (12.0)* | *230 (19.6)* |
| ***Education*** | | | | | | |
| < High School | 201 (15.1) | 198 (13.8) | 172 (14.1) | 230 (14.8) | 246 (16.9) | 228 (19.4) |
| Some High School | 592 (44.5) | 591 (41.2) | 470 (38.6) | 623 (40.1) | 573 (39.4) | 450 (38.3) |
| High School Grad | 387 (29.1) | 456 (31.8) | 402 (33.0) | 475 (30.6) | 446 (30.7) | 285 (24.3) |
| >= Some College | 111 (8.4) | 152 (10.6) | 143 (11.7) | 168 (10.8) | 119 (8.2) | 107 (9.1) |
| *Missing* | *38 (2.9)* | *39 (2.7)* | *31 (2.5)* | *57 (3.7)* | *71 (4.9)* | *104 (8.9)* |
| ***Hypertensive disorders of pregnancy*** | | | | | | |
| Normotensive | 1257 (94.6) | 1353 (94.2) | 1146 (94.1) | 1467 (94.5) | 1328 (91.3) | 1083 (92.2) |
| Chronic hypertension | 24 (1.8) | 20 (1.4) | 27 (2.2) | 32 (2.1) | 60 (4.1) | 41 (3.5) |
| Gestational hypertension | 23 (1.7) | 34 (2.4) | 28 (2.3) | 28 (1.8) | 37 (2.5) | 15 (1.3) |
| Preeclampsia/Eclampsia | 10 (0.8) | 13 (0.9) | 10 (0.8) | 9 (0.6) | 16 (1.1) | 16 (1.4) |
| Superimposed Preeclampsia/Eclampsia | 15 (1.1) | 15 (1.0) | 7 (0.6) | 17 (1.1) | 14 (1.0) | 11 (0.9) |
| *Missing* | *-* | *1 (0.1)* | *-* | *-* | *-* | *8 (0.7)* |
| ***Prior Diabetes*** ꝉ | | | | | | |
| No | 1307 (98.3) | 1424 (99.2) | 1198 (98.4) | 1527 (98.3) | 1421 (97.7) | 1123 (95.7) |
| Yes | 22 (1.7) | 12 (0.8) | 20 (1.6) | 26 (1.7) | 34 (2.3) | 44 (3.7) |
| *Missing* | *-* | *-* | *-* | *-* | *-* | *7 (0.6)* |
| ***Prior Cardiovascular Conditions*** ǂ | | | | | | |
| No | 1183 (89.0) | 1253 (87.3) | 1087 (89.2) | 1360 (87.6) | 1233 (84.7) | 1020 (86.9) |
| Yes | 145 (10.9) | 182 (12.7) | 131 (10.8) | 193 (12.4) | 222 (15.3) | 141 (12.0) |
| *Missing* | *1 (0.1)* | *1 (0.1)* | *-* | *-* | *-* | *13 (1.1)* |
| ***Prior Respiratory Conditions*** § | | | | | | |
| No | 1241 (93.4) | 1309 (91.2) | 1141 (93.7) | 1445 (93.0) | 1353 (93.0) | 1071 (91.2) |
| Yes | 87 (6.5) | 126 (8.8) | 77 (6.3) | 108 (7.0) | 102 (7.0) | 90 (7.7) |
| *Missing* | *1 (0.1)* | *1 (0.1)* | *-* | *-* | *-* | *13 (1.1)* |
| ***Prior Renal Conditions*** ¶ | | | | | | |
| No | 1269 (95.5) | 1382 (96.2) | 1176 (96.6) | 1477 (95.1) | 1407 (96.7) | 1102 (93.9) |
| Yes | 59 (4.4) | 53 (3.7) | 42 (3.4) | 76 (4.9) | 48 (3.3) | 59 (5.0) |
| *Missing* | *1 (0.1)* | *1 (0.1)* | *-* | *-* | *-* | *13 (1.1)* |
| ***Prior Neurological Conditions*** \|\| | | | | | | |
| No | 1220 (91.8) | 1330 (92.6) | 1122 (92.1) | 1445 (93.0) | 1340 (92.1) | 1061 (90.4) |
| Yes | 108 (8.1) | 105 (7.3) | 96 (7.9) | 108 (7.0) | 115 (7.9) | 100 (8.5) |
| *Missing* | *1 (0.1)* | *1 (0.1)* | *-* | *-* | *-* | *13 (1.1)* |
| ***Prior Cancer or Tumors*** ** | | | | | | |
| No | 1287 (96.8) | 1375 (95.8) | 1182 (97.0) | 1508 (97.1) | 1404 (96.5) | 1136 (96.8) |
| Yes | 41 (3.1) | 60 (4.2) | 36 (3.0) | 45 (2.9) | 51 (3.5) | 25 (2.1) |
| *Missing* | *1 (0.1)* | *1 (0.1)* | *-* | *-* | *-* | *13 (1.1)* |
| ***Pregnancy Plurality*** | | | | | | |
| Singleton | 1314 (98.9) | 1423 (99.1) | 1214 (99.7) | 1541 (99.2) | 1443 (99.2) | 1125 (95.8) |
| Multiples | 13 (1.0) | 11 (0.8) | 4 (0.3) | 12 (0.8) | 11 (0.8) | 11 (0.9) |
| *Missing* | *2 (0.2)* | *2 (0.1)* | *0 (0.0)* | *0 (0.0)* | *1 (0.1)* | *38 (3.2)* |
| ***Study Site*** | | | | | | |
| Boston | 303 (22.8) | 418 (29.1) | 429 (35.2) | 522 (33.6) | 461 (31.7) | 340 (29.0) |
| Buffalo | 60 (4.5) | 74 (5.2) | 61 (5.0) | 66 (4.2) | 50 (3.4) | 30 (2.6) |
| New Orleans | 22 (1.7) | 24 (1.7) | 18 (1.5) | 32 (2.1) | 40 (2.7) | 5 (0.4) |
| New York/Columbia | 17 (1.3) | 12 (0.8) | 8 (0.7) | 14 (0.9) | 10 (0.7) | 9 (0.8) |
| Baltimore | 78 (5.9) | 97 (6.8) | 77 (6.3) | 131 (8.4) | 120 (8.2) | 88 (7.5) |
| Virginia | 54 (4.1) | 82 (5.7) | 50 (4.1) | 88 (5.7) | 108 (7.4) | 93 (7.9) |
| Minnesota | 97 (7.3) | 103 (7.2) | 62 (5.1) | 55 (3.5) | 29 (2.0) | 41 (3.5) |
| New York/Medical College | 40 (3.0) | 33 (2.3) | 43 (3.5) | 40 (2.6) | 48 (3.3) | 72 (6.1) |
| Oregon | 159 (12.0) | 108 (7.5) | 66 (5.4) | 74 (4.8) | 80 (5.5) | 60 (5.1) |
| Pennsylvania | 356 (26.8) | 318 (22.1) | 274 (22.5) | 340 (21.9) | 334 (23.0) | 345 (29.4) |
| Providence | 118 (8.9) | 128 (8.9) | 96 (7.9) | 134 (8.6) | 106 (7.3) | 77 (6.6) |
| Tennessee | 25 (1.9) | 39 (2.7) | 34 (2.8) | 57 (3.7) | 69 (4.7) | 14 (1.2) |
| *Missing* | *-* | *-* | *-* | *-* | *-* | *-* |
| ***Pregnancy outcome*** | | | | | | |
| 0: liveborn | 1279 (96.2) | 1388 (96.7) | 1159 (95.2) | 1459 (93.9) | 1185 (81.4) | 971 (82.7) |
| 1: pregnancy loss | 11 (0.8) | 15 (1.0) | 17 (1.4) | 45 (2.9) | 196 (13.5) | 120 (10.2) |
| 2: neonatal death | 14 (1.1) | 11 (0.8) | 21 (1.7) | 35 (2.3) | 55 (3.8) | 29 (2.5) |
| 3: infant/child death | 23 (1.7) | 20 (1.4) | 21 (1.7) | 14 (0.9) | 18 (1.2) | 16 (1.4) |
| *Missing* | 2 (0.2) | 2 (0.1) | 0 (0.0) | 0 (0.0) | 1 (0.1) | 38 (3.2) |
| ***Height (cm)*** | | | | | | |
| Mean (SD) | 162.0 (6.1) | 161.2 (6.0) | 160.6 (6.3) | 160.5 (6.3) | 160.7 (6.5) | 160.5 (6.8) |
| *Missing* | *15 (1.1)* | *12 (0.8)* | *15 (1.2)* | *15 (1.0)* | *11 (0.8)* | *72 (6.1)* |
| ***Pre-pregnancy Weight (kg)*** | | | | | | |
| Mean (SD) | 59.2 (10.8) | 56.4 (8.8) | 56.6 (9.0) | 57.2 (10.0) | 61.6 (12.9) | 60.2 (12.6) |
| *Missing* | *-* | *-* | *-* | *-* | *-* | *131 (11.2)* |
| ***Pre-pregnancy BMI (kg/m2)*** | | | | | | |
| Mean (SD) | 22.5 (3.9) | 21.7 (3.1) | 21.9 (3.2) | 22.2 (3.6) | 23.8 (4.7) | 23.4 (4.6) |
| Median (IQR) | 21.9 (19.9-24.2) | 21.1 (19.6-23.2) | 21.3 (19.8-23.2) | 21.5 (19.7-23.9) | 22.8 (20.5-26.1) | 22.3 (20.3-25.0) |
| *Missing* | *15 (1.1)* | *12 (0.8)* | *15 (1.2)* | *15 (1.0)* | *11 (0.8)* | *173 (14.7)* |
| ***Pre-pregnancy BMI (kg/m2) categories*** | | | | | | |
| Underweight: < 18.5 kg/m2 | 126 (9.5) | 162 (11.3) | 105 (8.6) | 154 (9.9) | 95 (6.5) | 88 (7.5) |
| Normal Weight: 18.5 – 24.9 kg/m2 | 923 (69.5) | 1087 (75.7) | 919 (75.5) | 1118 (72.0) | 876 (60.2) | 656 (55.9) |
| Overweight: 25.0 – 29.9 kg/m2 | 197 (14.8) | 146 (10.2) | 145 (11.9) | 210 (13.5) | 317 (21.8) | 163 (13.9) |
| Obese: > 30 kg/m2 | 68 (5.1) | 29 (2.0) | 34 (2.8) | 56 (3.6) | 156 (10.7) | 94 (8.0) |
| *Missing* | *15 (1.1)* | *12 (0.8)* | *15 (1.2)* | *15 (1.0)* | *11 (0.8)* | *173 (14.7)* |
| ***Birth Weight (g)*** | | | | | | |
| Mean (SD) | 3373 (545.3) | 3206 (517.3) | 3125 (593.4) | 3060 (640.9) | 2886 (886.2) | 3050 (803.0) |
| *Missing* | *6 (0.5)* | *5 (0.3)* | *5 (0.4)* | *12 (0.8)* | *111 (7.6)* | *121 (10.3)* |
| ***Gestational Weight Gain (kg)*** | | | | | | |
| Mean (SD) | 13.8 (4.5) | 10.9 (3.3) | 9.4 (3.4) | 8.3 (3.7) | 6.3 (4.8) | 9.2 (4.8) |
| *Missing* | *-* | *-* | *-* | *-* | *-* | *603 (51.4)* |
| ***Gestational Age at Delivery (weeks)*** | | | | | | |
| Mean (SD) | 40.0 (3.0) | 39.6 (3.2) | 39.2 (3.3) | 38.9 (3.9) | 36.0 (8.0) | 38.0 (6.1) |
| *Missing* | *7 (0.5)* | *5 (0.3)* | *2 (0.2)* | *4 (0.3)* | *7 (0.5)* | *17 (1.4)* |
| ***Interpregnancy Interval (in years)*** * | | | | | | |
| Mean (SD) | 1.2 (1.0) | 1.1 (0.9) | 1.1 (0.9) | 1.2 (1.0) | 1.4 (1.1) | 1.3 (1.1) |
| *Missing* | *2 (0.2)* | *1 (0.1)* | *-* | *1 (0.1)* | *1 (0.1)* | *668 (56.9)* |
| ꝉ Type unknown. ‡ Cardiovascular conditions included hypertension, rheumatic fever, and any other cardiovascular diseases. §Respiratory conditions included tuberculosis, asthma, other chronic pulmonary diseases, and other conditions requiring thoracic surgery. ¶ Renal conditions included pyelitis, glomerulonephritis, and other conditions requiring kidney, urinary, or bladder surgery. \|\|Neurological conditions included neuromuscular diseases, convulsive disorders psychosis, alcohol or drug addiction, or other neurological diseases. **Cancer and tumors included any history of cancer, and gastrointestinal, kidney, urinary, bladder, or gynecological tumors.  * Interpregnancy Interval calculated as the time difference between 1st pregnancy delivery date and LMP of the 2nd pregnancy | | | | | | |

**Supplementary Table 5**: Distribution of outcome of interest across quintiles of interpregnancy weight change (IPWC)

| **Outcomes of Interest** | | **IPWC quintile** | | | | | |
| --- | --- | --- | --- | --- | --- | --- | --- |
|  |  | **Q1: < -11.8 kg (n = 1329)** | **Q2: ≥-11.8 to <-9.1 kg (n = 1436)** | **Q3: ≥-9.1 to <-7.3 kg (n = 1218)** | **Q4: ≥-7.3 to <-4.5 kg (n = 1553)** | **Q5: ≥-4.5 kg (n = 1455)** | **Missing (n = 1174)** |
|  |  | **n (%)** | **n (%)** | **n (%)** | **n (%)** | **n (%)** | **n (%)** |
| Vital Status | Alive | 771 (58.0) | 900 (62.7) | 737 (60.5) | 926 (59.6) | 767 (52.7) | 659 (56.1) |
|  | Dead | 558 (42.0) | 536 (37.3) | 481 (39.5) | 627 (40.4) | 688 (47.3) | 515 (43.9) |
|  | *Missing* | *0 (0.0)* | *0 (0.0)* | *0 (0.0)* | *0 (0.0)* | *0 (0.0)* | *0 (0.0)* |
| Cause of Death | Cardiovascular | 158 (28.3) | 124 (23.1) | 105 (21.8) | 166 (26.5) | 199 (28.9) | 147 (28.5) |
|  | Diabetes | 27 (4.8) | 19 (3.5) | 14 (2.9) | 19 (3.0) | 25 (3.6) | 32 (6.2) |
|  | Kidney | 10 (1.8) | 2 (0.4) | 11 (2.3) | 7 (1.1) | 20 (2.9) | 9 (1.7) |
|  | *Missing* | *47 (3.5)* | *63 (4.4)* | *52 (4.3)* | *59 (3.8)* | *51 (3.5)* | *53 (4.5)* |

**Supplementary Figure 5:** Risk of all cause mortality associated with quintiles of interpregnancy weight change (IPWC) with weight gain group seperated out

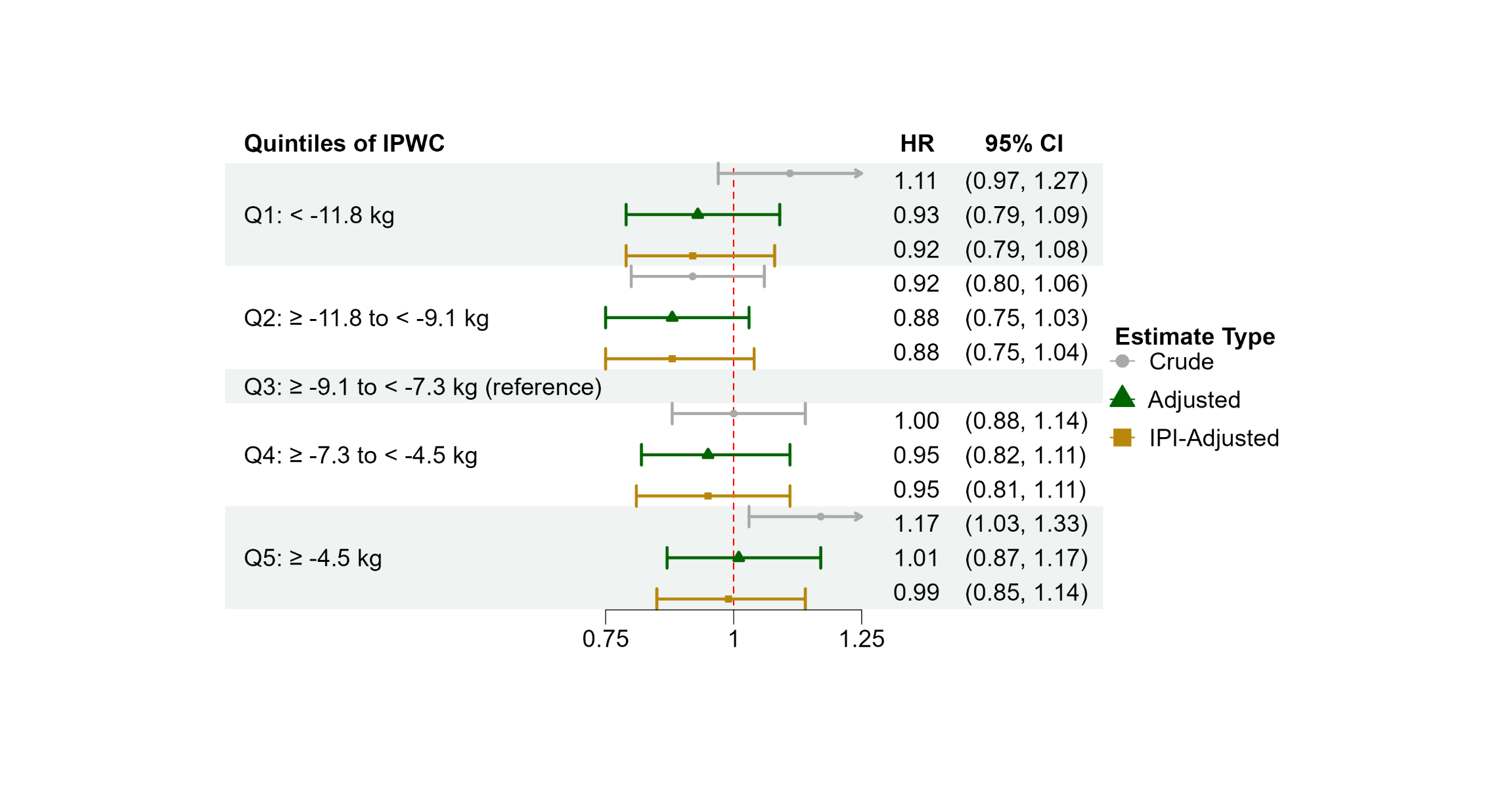

IPI: Interpregnancy interval

Adjusted for CPP 1^st^ Pregnancy variables: age, race and ethnicity, number of prior pregnancies, marital status, smoking status, annual income, education, hypertensive disorders of pregnancy, prior diabetes, prior cardiovascular conditions, prior respiratory conditions, prior renal conditions, prior neurological conditions, prior cancer or tumors, pregnancy plurality, study site, pre-pregnancy BMI, and gestational weight gain

**Supplementary Table 6:** All-cause mortality risks associated with BMI-specific quintiles of interpregnancy weight change (IPWC), and quintiles of IPWC with weight gain separated out for those with Normal pre-pregnancy BMI

| **Exposure** | **Category (in kg)** | **Unadjusted** | **Adjusted *** | |
| --- | --- | --- | --- | --- |
|  |  | **HR (95% CI)** | **HR (95% CI) adjusted without IPI** | **HR (95% CI) adjusted with IPI** |
| **BMI-specific quintiles of IPWC** | Q1: ≤ -11.8 | 1.18 (1.03-1.35) | 0.98 (0.84-1.14) | 0.97 (0.83-1.13) |
|  | Q2: > -11.8 to -9.5 | 1.02 (0.88-1.19) | 0.96 (0.82-1.14) | 0.97 (0.82-1.14) |
|  | Q3: > -9.5 to -7.3 | reference | | |
|  | Q4: > -7.3 to -5.0 | 1.15 (0.99-1.34) | 1.13 (0.96-1.33) | 1.12 (0.95-1.31) |
|  | Q5: > -5.0 | 1.17 (1.02-1.35) | 1.21 (1.02-1.43) | 1.17 (0.99-1.39) |
| **BMI-specific quintiles of IPWC with weight gain separated out** | 1: ≤ -11.8 | 1.18 (1.03-1.35) | 0.98 (0.84-1.14) | 0.97 (0.83-1.13) |
|  | 2: > -11.8 to -9.5 | 1.02 (0.88-1.19) | 0.96 (0.81-1.14) | 0.97 (0.82-1.14) |
|  | 3: > -9.5 to -7.3 | reference | | |
|  | 4: > -7.3 to -5.0 | 1.15 (0.99-1.34) | 1.13 (0.96-1.33) | 1.12 (0.95-1.31) |
|  | 5: > -5.0 to < 0 | 1.13 (0.96-1.32) | 1.17 (0.98-1.38) | 1.14 (0.96-1.35) |
|  | 6: ≥ 0 | 1.34 (1.06-1.69) | 1.35 (1.01-1.79) | 1.29 (0.98-1.71) |
| * Adjusted for CPP 1^st^ Pregnancy variables: age, race and ethnicity, number of prior pregnancies, marital status, smoking status, annual income, education, hypertensive disorders of pregnancy, prior diabetes, prior cardiovascular conditions, prior respiratory conditions, prior renal conditions, prior neurological conditions, prior cancer or tumors, pregnancy plurality, study site, and gestational weight gain | | | | |

**Supplementary Table 7:** All-cause mortality risks associated with quintiles of interpregnancy weight change (IPWC), and quintiles of IPWC with weight gain separated out adjusting for birth weight and gestational age at delivery

| **Exposure** | **Category (in kg)** | **Birth weight-Adjusted *** | | **Gestational age at delivery-Adjusted *** | | **Pregnancy Outcome-Adjusted *** | |
| --- | --- | --- | --- | --- | --- | --- | --- |
|  |  | **HR (95% CI) adjusted without IPI** | **HR (95% CI) adjusted with IPI** | **HR (95% CI) adjusted without IPI** | **HR (95% CI) adjusted with IPI** | **HR (95% CI) adjusted without IPI** | **HR (95% CI) adjusted with IPI** |
| **Quintiles of IPWC** | Q1: < -11.8 | 0.93 (0.79-1.09) | 0.92 (0.79-1.08) | 0.93 (0.79-1.09) | 0.92 (0.79-1.08) | 0.93 (0.79-1.09) | 0.92 (0.79-1.08) |
|  | Q2: ≥ -11.8 to < -9.1 | 0.88 (0.75-1.03) | 0.88 (0.75-1.04) | 0.88 (0.75-1.03) | 0.88 (0.75-1.04) | 0.88 (0.75-1.03) | 0.88 (0.75-1.04) |
|  | Q3: > -9.1 to -7.3 | reference | | | | | |
|  | Q4: ≥ -7.3 to < -4.5 | 0.95 (0.82-1.11) | 0.95 (0.81-1.10) | 0.95 (0.82-1.11) | 0.95 (0.81-1.11) | 0.96 (0.82-1.11) | 0.95 (0.81-1.11) |
|  | Q5: ≥ -4.5 | 0.99 (0.86-1.15) | 0.97 (0.84-1.13) | 1.01 (0.87-1.17) | 0.99 (0.85-1.15) | 1.01 (0.87-1.17) | 0.99 (0.85-1.14) |
| **Quintiles of IPWC with weight gain separated out** | 1: < -11.8 | 0.92 (0.79-1.09) | 0.92 (0.79-1.08) | 0.93 (0.79-1.09) | 0.92 (0.79-1.08) | 0.93 (0.79-1.09) | 0.92 (0.79-1.08) |
|  | 2: ≥ -11.8 to < -9.1 | 0.88 (0.75-1.03) | 0.88 (0.75-1.04) | 0.88 (0.75-1.03) | 0.88 (0.75-1.04) | 0.88 (0.75-1.03) | 0.88 (0.75-1.04) |
|  | 3: > -9.1 to -7.3 | reference | | | | | |
|  | 4: ≥ -7.3 to < -4.5 | 0.95 (0.82-1.11) | 0.95 (0.81-1.11) | 0.96 (0.82-1.12) | 0.95 (0.81-1.11) | 0.96 (0.82-1.12) | 0.95 (0.82-1.11) |
|  | 5: ≥ -4.5 | 0.95 (0.81-1.10) | 0.93 (0.80-1.08) | 0.96 (0.82-1.12) | 0.94 (0.81-1.10) | 0.96 (0.82-1.12) | 0.94 (0.81-1.10) |
|  | 6: ≥ 0 | 1.14 (0.93-1.40) | 1.10 (0.90-1.35) | 1.17 (0.95-1.44) | 1.13 (0.92-1.39) | 1.17 (0.95-1.43) | 1.13 (0.92-1.38) |
| * Adjusted for CPP 1^st^ Pregnancy variables: age, race and ethnicity, number of prior pregnancies, marital status, smoking status, annual income, education, hypertensive disorders of pregnancy, prior diabetes, prior cardiovascular conditions, prior respiratory conditions, prior renal conditions, prior neurological conditions, prior cancer or tumors, pregnancy plurality, study site, pre-pregnancy BMI, and gestational weight gain | | | | | | | |

**Supplementary Table 8:** Cause-specific mortality associated with quintiles of interpregnancy weight change (IPWC)

| **Cause of Death** | **Exposure (quintiles of IPWC in kg)** | **Unadjusted** | **Adjusted *** | |
| --- | --- | --- | --- | --- |
|  |  | **HR (95% CI)** | **HR (95% CI) adjusted without IPI** | **HR (95% CI) adjusted with IPI** |
| **Cardiovascular** | Q1: < -11.8 | 1.25 (0.88-1.77) | 1.02 (0.69-1.51) | 1.01 (0.69-1.49) |
|  | Q2: ≥ -11.8 to < -9.1 | 0.99 (0.72-1.35) | 0.98 (0.69-1.38) | 0.98 (0.69-1.38) |
|  | Q3: > -9.1 to -7.3 | reference | | |
|  | Q4: ≥ -7.3 to < -4.5 | 1.19 (0.88-1.60) | 1.15 (0.83-1.60) | 1.14 (0.83-1.58) |
|  | Q5: ≥ -4.5 | 1.38 (0.98-1.94) | 1.11 (0.78-1.57) | 1.08 (0.76-1.53) |
| **Diabetes** | Q1: < -11.8 | 1.68 (0.95-2.95) | 0.87 (0.45-1.70) | 0.87 (0.45-1.69) |
|  | Q2: ≥ -11.8 to < -9.1 | 0.83 (0.44-1.57) | 0.85 (0.42-1.69) | 0.86 (0.43-1.71) |
|  | Q3: > -9.1 to -7.3 | reference | | |
|  | Q4: ≥ -7.3 to < -4.5 | 1.07 (0.58-1.98) | 1.13 (0.60-2.14) | 1.11 (0.59-2.11) |
|  | Q5: ≥ -4.5 | 1.26 (0.75-2.13) | 1.05 (0.59-1.86) | 1.01 (0.56-1.81) |
| **Kidney** | Q1: < -11.8 | 1.05 (0.42-2.64) | 0.72 (0.26-2.01) | 0.72 (0.26-1.99) |
|  | Q2: ≥ -11.8 to < -9.1 | 0.42 (0.09-1.95) | 0.37 (0.08-1.75) | 0.38 (0.08-1.76) |
|  | Q3: > -9.1 to -7.3 | reference | | |
|  | Q4: ≥ -7.3 to < -4.5 | 0.80 (0.25-2.51) | 0.80 (0.24-2.65) | 0.80 (0.24-2.61) |
|  | Q5: ≥ -4.5 | 2.12 (0.89-5.04) | 1.75 (0.68-4.47) | 1.72 (0.68-4.37) |
| * Adjusted for CPP 1^st^ Pregnancy variables: age, race and ethnicity, number of prior pregnancies, marital status, smoking status, annual income, education, hypertensive disorders of pregnancy, prior diabetes, prior cardiovascular conditions, prior respiratory conditions, prior renal conditions, prior neurological conditions, prior cancer or tumors, pregnancy plurality, study site, pre-pregnancy BMI, and gestational weight gain | | | | |
